# Multi-omic landscaping of human midbrains identifies neuroinflammation as major disease mechanism in advanced-stage Parkinson’s disease

**DOI:** 10.1101/2021.06.08.21258527

**Authors:** Lucas Caldi Gomes, Ana Galhoz, Gaurav Jain, Anna-Elisa Roser, Fabian Maass, Eleonora Carboni, Elisabeth Barski, Christof Lenz, Katja Lohmann, Christine Klein, Mathias Bähr, André Fischer, Michael P. Menden, Paul Lingor

## Abstract

Parkinson’s disease (PD) is the second most common neurodegenerative disorder whose prevalence is rapidly increasing worldwide. The disease mechanisms of sporadic PD are not yet completely understood. Therefore, causative therapies are still lacking. To obtain a more integrative view of disease-mediated alterations, we investigated the molecular landscape of PD in human post-mortem midbrains. Tissue from 13 PD patients and 10 controls was subjected to small RNA sequencing, transcriptomics, and proteomics analysis. Differential expression analyses were performed reveal multiple deregulated molecular targets linked to known pathomechanisms of PD as well as novel processes. We found significant differential expression of miR-539-3p, miR-376a-5p, miR-218-5p, and miR-369-3p, the valid miRNA-mRNA interacting pairs of miR-218-5p/*RAB6C*, and miR-369-3p/*GTF2H3*, as well as multiple proteins relevant in the pathology of PD, including CHI3L1, SELENBP1, PRDX1, HSPA1B, and TH. Vertical integration of multiple omics analyses allowed to validate disease-mediated molecular alterations across different molecular layers and functional annotation of differentially expressed targets identified a strong enrichment of pathways related to inflammation and activation of the immune response. This suggests that neuroinflammation may significantly contribute to disease progression in PD and may be a promising therapeutic target in advanced stages of PD.

## Main Text

### Introduction

Parkinson’s Disease (PD) is the fastest-growing neurodegenerative disorder and affects up to 2% of individuals aged over 60 years (1). While the chronic and progressive motor dysfunction is mostly due to degeneration of dopaminergic neurons in the nigrostriatal pathway, PD is now recognized to be a systemic disorder involving multiple other regions of the nervous system (2). The sporadic form accounts for most cases of PD, whereas only up to 3% of cases comprise the autosomal forms (3). Several environmental and genetic factors are known to increase the disease risk (4). Recent studies point to a multifactorial pathogenesis that may differ between patients, suggesting that PD is not one homogeneous disease entity, but rather a syndrome with a unifying clinical phenotype and numerous molecular subgroups. The existence of non-motor symptoms that appear many years before the onset of motor manifestations suggest that molecular mechanisms may have a long lead-up period and result in chronic degeneration (5). One of the pathological hallmarks of PD is the presence of Lewy-bodies (LB), intracytoplasmic inclusions that are majorly composed of the protein alpha-synuclein (αSyn), but also contain ubiquitin and neurofilaments (6). Such proteinaceous inclusions occur in both familial and sporadic forms of PD, suggesting that defects in the protein handling machinery are directly related to the pathogenesis of the disease (7).

Previously, neuroinflammatory mechanisms have been linked to PD, although their contribution remains unclear. For example, activated microglia have been localized in the substantia nigra of brains of patients with PD (8) as well as lymphocytic infiltration and astrogliosis (9). A chronic inflammatory state could contribute to the pathogenesis through the release of neurotoxic cytokines by glial cells, which further promotes neuronal cell damage (10). Our current understanding of disease progression also considers the involvement of different disease mechanisms in different stages of the disease. Environmental factors are likely to represent triggers that start a pathological cascade of events that lead to the facilitation of molecular alterations which are further aggravated as the disease progresses (5). Furthermore, mechanisms related to the regulation of gene expression have been extensively linked to the development and progression of a variety of brain diseases in the last years. For instance, alterations of miRNA expression have been linked to the development and progression of a variety of brain diseases, including PD (11), where deregulated miRNAs have been identified in nervous system tissues and in peripheral fluids (12, 13). Decreased levels of miR-133b were identified in the midbrain of patients with PD and in mouse models of PD (14, 15). Alterations in the levels of miR-34b/c were found in several regions of PD-affected brains. These miRNAs can mimic impairments in mitochondrial function and oxidative stress, disease mechanisms believed to be crucial for the development of PD (16). Two miRNAs (miR-7 and miR-153) were also shown to regulate the expression of αSyn. Interestingly, the former has been found to be altered in the striatum and substantia nigra of patients with PD, as well as in murine models of PD (17, 18). Exploring individual profiles of the transcriptome, the microRNAome, or the proteome in PD-affected brains is a powerful strategy to understand neurodegenerative events that underlie the disease. For instance, defects in iron metabolism (19), in autophagy (20), mitochondrial dysfunctions (21), and dysfunctions in synaptic function (19, 22) were all captured in previous transcriptomic and/or proteomics studies. Moreover, a number of promising targets were identified by such studies and have been regarded as potential therapeutic targets or disease biomarkers for PD (e.g. NR4A2, ULK1, OR51E2, NRF2, FTL, GGH, BSCL2)(19–24). Nevertheless, singular omic profiling also encompasses a number of limitations, as it provides only a snapshot of the pathological events that might be a part of a much bigger network of deregulation, and fails to capture changes occurring up-/down-stream to the selected omic level (e.g. miRNA regulation or post-translational modifications). Only few studies have attempted to combine high-throughput profiling techniques to explore the molecular changes that take place in postmortem PD-affected brains, but the number of techniques employed in parallel was limited (25–28). A comprehensive and integrative assessment of postmortem PD-affected brains across genomic, miRNAomic, transcriptomic, and proteomic levels has not been performed so far. Therefore, in this study, we analyzed DNA, RNA, and proteins of postmortem midbrain tissue from a cohort of PD patients and controls (CTR) to obtain a multi-omic landscape of the disease. Each molecular layer was analyzed individually and in an integrative fashion, aiming to depict deregulated pathways that permit the exploration of molecular changes taking place in PD-affected brains. Overall, our findings point to putative molecular networks involved in the pathophysiology of PD, which might improve disease monitoring and delineate novel druggable targets for this devastating disease

## Results

In the present analysis, we leveraged a comprehensive multi-omics dataset of a cohort of midbrain samples including those of patients with PD and those of individuals without any indication of neurodegeneration (CTR). We obtained human midbrain tissue samples from 19 PD and 12 CTR individuals from the Parkinson’s UK Brain Bank in two batches. The largest batch comprising 13 PD and 10 CTR cases was used as the discovery cohort and processed to obtain protein, RNA, and DNA lysates, which were further subjected to a multi-omic analysis (**Fig.1A**). Our discovery cohort dataset comprised 57,992 total RNA, 31,186 small RNA (of which 4,383 are miRNAs) and 2,257 proteins (**Fig.1B**). The second batch, 6 PD and 2 CTR cases, was used for validation.

**Fig. 1.**
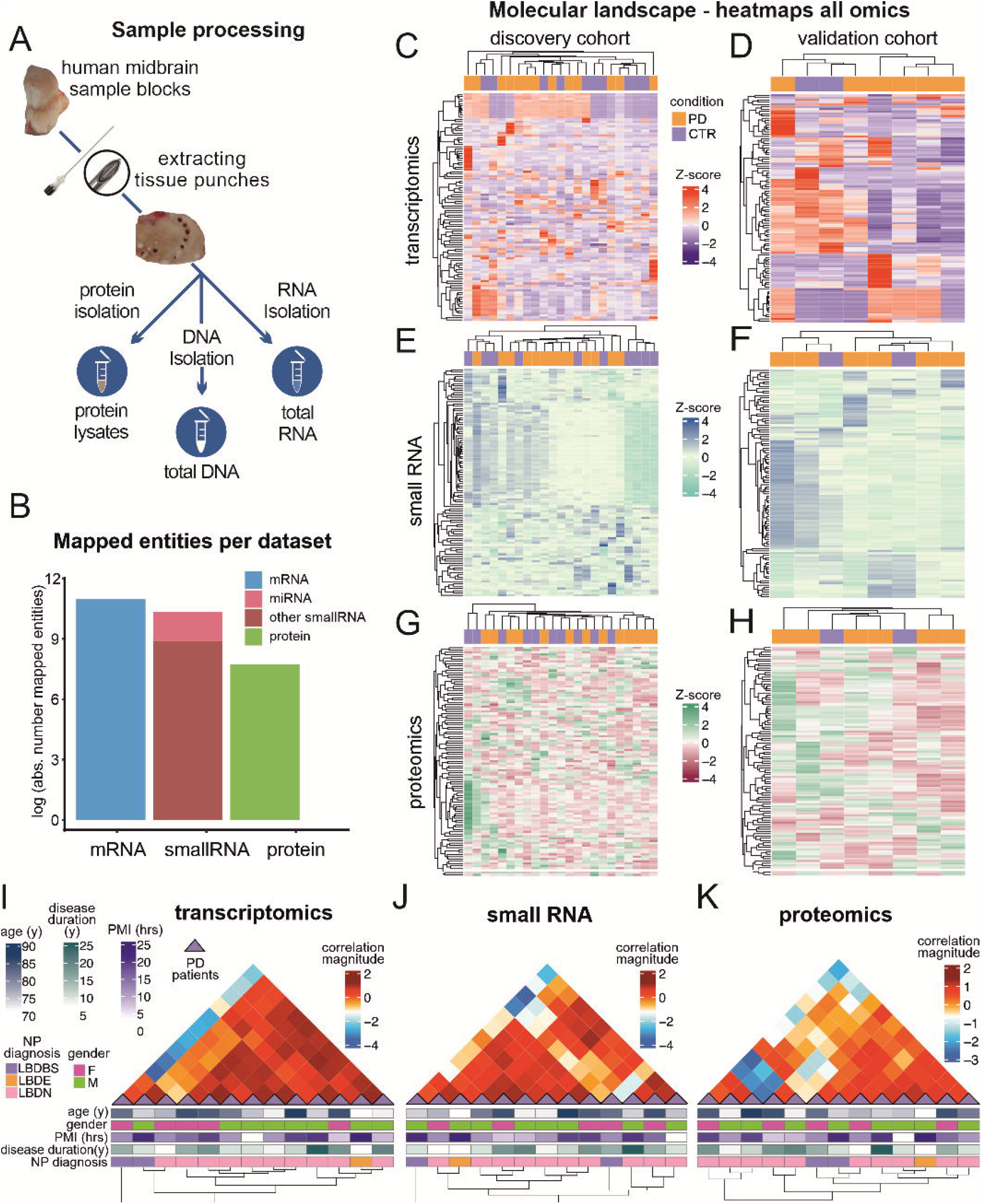
Overview of multi-omics profiles. (A) Experimental design for the isolation of RNA, DNA and proteins from human midbrain sample blocks. The extraction of tissue biopsies was performed with a spinal needle. Adjacent tissue biopsies from each sample were used for the different isolation techniques. (B) Barplot representing the number of mapped entities for each omics dataset. The y-axis represents the natural log of the mapped entities. (C)-(H) Heatmaps of the top 100 most variant transcripts, small RNAs and proteins for the validation and discovery cohort. The diagrams display the z-score computed from the normalized counts for each individual. Column dendrograms were obtained based on the selected omics’ molecular profiles, and the row groups depict the samples’ effect. Both clusters were determined using Euclidean distance and a complete hierarchical clustering. (I-K) Unbiased Bayesian hierarchical clustering of PD samples according to the total and small RNA, and proteomics expression profiles. Clinical parameters for each PD patient are represented in the lower panel of the illustrations. The column dendrograms depict the unsupervised clustering based on the correlation between patients.

### Multi-omics expression patterns and Bayesian hierarchical clustering analyses

Samples were hierarchically grouped according to the level of expression of each of the omics datasets (**Fig.1C-H**). The 100 most variable transcripts, small RNAs, and proteins across the discovery and validation cohorts resulted in mixed sample clusters between PD and CTR, indicating a high expression diversity in both groups. A Bayesian Hierarchical Clustering of the normalized mapped counts of transcriptomics, small RNA, and proteomics expression data in the PD group only showed different levels of heterogeneity with a decreasing level of positive correlation between subjects, starting from the transcriptome over the small RNA composition up to the proteome (with 64, 61, and 57 positively correlated pairs of patients for transcriptomics, small RNA, and proteomics, respectively). Although sub-clusters of expression patterns with a higher correlation magnitude were discernible, they did not correlate with patterns of respective clinical/histological parameters, suggesting that molecular diversity in PD may be independent of the clinical/histological phenotype (**Fig.1I-K**; **Supplementary Methods**).

### Genetic background of PD patients

To describe the genetic background of PD patients that were selected for this study and to exclude bias from mutations known to cause familial PD, we performed both gene panel sequencing and Multiplex Ligation-dependent Probe Amplification (MLPA)(**Fig.S1**). Here, the presence of mutations/duplications/triplications in genes previously associated with PD was assessed. MLPA results revealed no alterations in copy numbers in any of the analyzed genes/exons (**Fig.S1B**;**Table S1**). A panel of 29 genes/exons previously linked to PD or dystonia (DYT) phenotypes (**Fig.S1C**;**Table S2**) was employed for targeted next-generation sequencing analysis of the PD patient cohort. No pathogenic or likely pathogenic variants were identified using gene panel sequencing. One patient (PD6) presented a variant of uncertain significance (VUS) for the *POLG* gene (single nucleotide variant, NM_001126131.1:c. 2542G>A).

### Expression profiles of small and total RNA, functional annotation and integration

After isolation of total RNA, we analyzed the small RNA content and profiled miRNA expression patterns using small and total RNA sequencing (**Fig.S2A**). To analyze differential expression (DE) of total and small RNA, we leveraged two bioinformatics frameworks, which we call “A” and “B” (**Fig.2A**). These frameworks differed in the pre-processing stage: “A” was data-driven and performed filtering using the distribution quantile information, while “B” took a more supervised approach and removed the negative control omics (i.e., omics with zero effect on the variance)(**Supplementary methods**). Following small RNA sequencing, mapping results showed that most sequencing counts were composed of miRNAs in both PD and CTR conditions (90.39% and 92.61% of all mapped counts for different small RNA species, respectively)(**Fig.3B**). Using the same RNA source, mRNA libraries were prepared using a strand-specific, massive-parallel cDNA library preparation protocol. Sequencing read counts were mapped and assigned to the reference genome, accounting for a total of 46,500 genes with valid read values before filtering. For both frameworks, data quality assessments were conducted using unsupervised learning methods (**Fig.S3B-F**). Principal component analysis of the small RNA seq data revealed that the control (CTR12) did not cluster with the rest of the samples (**Fig.S3D**). The retrieved neuropathological report for this sample indicated the presence of AD-type Tau pathology-Braak-stage II (29), and multiple small demyelinating plaques, which also suggested that it was a potential asymptomatic multiple sclerosis case (30). Following an outlier comparative analysis via the Grubbs-test (p-value=0.033), this sample was regarded as an outlier and removed from further analyses (**Fig.S3E**). Based on p-adjusted<0.1 and log_2_FC>0 criteria, we defined significantly different genes and miRNAs for both frameworks. For framework “A”, DE analyses revealed 641 regulated genes (512 up- and 129 down-regulated in PD) but no DE miRNAs (**Fig.2B**,**3B**,**3D;Supplementary dataset 1**). Application of framework “B” resulted in 126 DE genes (105 up- and 21 down-regulated in PD) and 4 significantly DE miRNAs (miR-539-3p; miR-376a-5p; miR-218-5p; miR-369-3p) all of which were up-regulated in PD (**Fig.2C**,**3B**,**3E**;**Supplementary dataset 2**). In addition, the frameworks shared 101 common genes, all with the same regulation directionality regarding PD and CTR. Our results demonstrate the usage of two analysis pipelines for one data set encompassing multiple characteristics: framework “A” was suitable for diverse data with high variability between biological groups, such as our gene expression data (**Fig.S4A**), whereas framework “B” was more sensitive to omic data with low expression and less dispersion, which is true for our miRNA expression data (**Fig.S4B**).

**Fig. 2.**
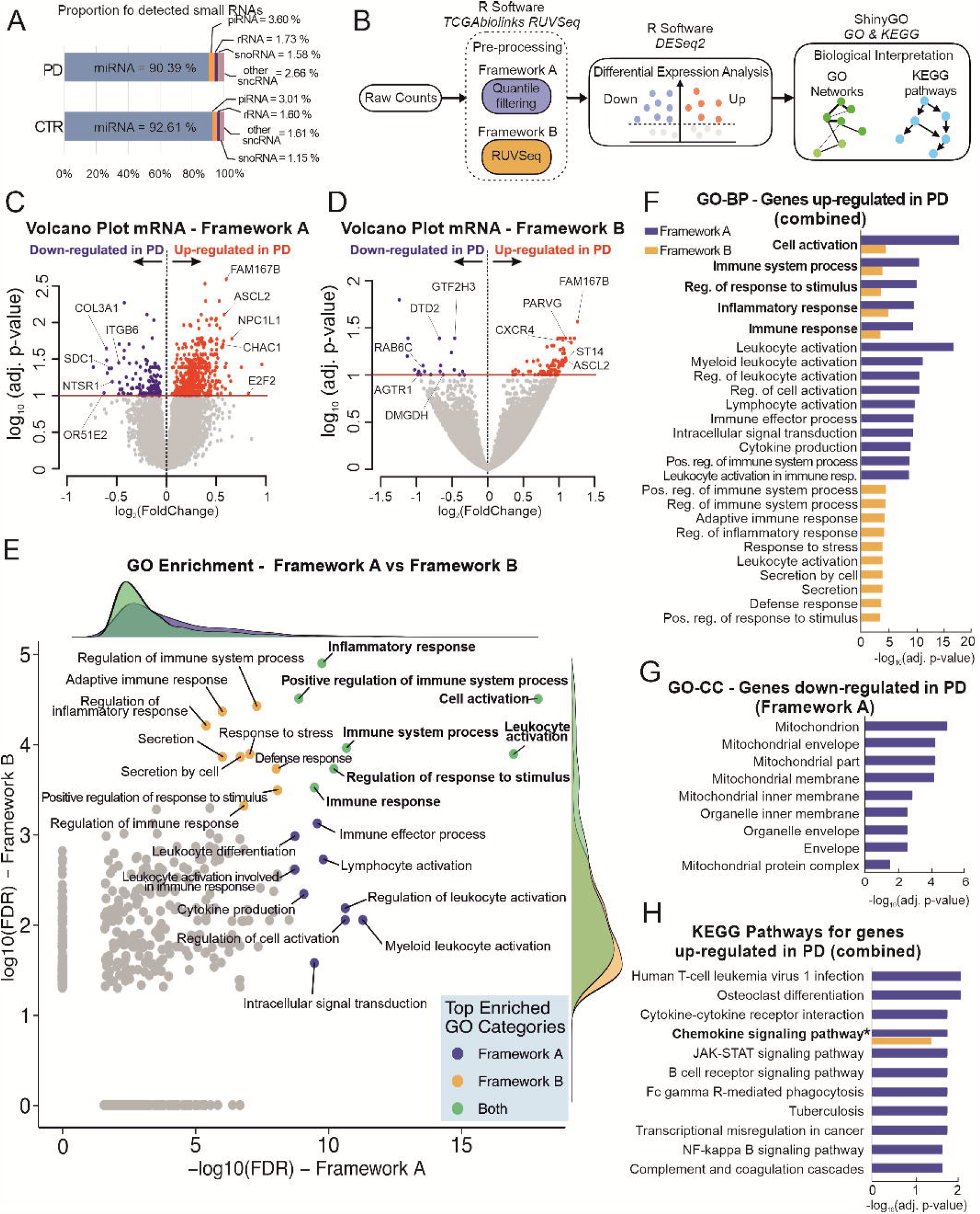
RNA-seq data schematic workflow and analysis results for total RNA-seq. (A) Illustration of the workflow of the bioinformatics pipelines used for differential expression analysis of small and total RNA sequencing data. The analyses start with the raw data expressed as integer reads for each sample and small RNA/gene. These were pre-processed using two distinct frameworks “A” and “B” (details in Supplementary Information). Then, the frameworks were further evaluated for differential expression and functional annotation. (B and C) Volcano plots portraying the differential expression of total RNA sequencing data between PD and CTR subjects, for frameworks “A” and “B”, respectively. The x-axis represents log2(fold change) (log_2_FC) and y-axis -log10(p-adjusted value). Under p-adjusted<0.1 and |log_2_FC|>0, we found 641 and 126 differentially expressed genes for framework “A” and “B”, respectively. Genes attending these criteria are colored in blue and red, for negative and positive log_2_FC, respectively. Highlighted genes based on the integrative analyses for RNA sequencing experiments. (D) Comparison of enriched false discovery rate (FDR) gene ontology (GO) categories obtained by frameworks “A” and “B” for the significantly up-regulated genes (under p-value cut-off of 0.05, yielding 500 and 427 enriched GO categories for framework “A” and “B”, respectively). Only commonly enriched categories were considered for the scatterplot. The top enriched GO categories are highlighted for framework “A” (-log10(FDR)>8.7, a total of 16 classes, in blue), “B” (-log10(FDR)>3.3, a total of 15 classes, in orange) and both (in green). Marginal plots represent densities of enriched GO classes for each framework and ensemble. The axis values are in the base-10 log scale. Additionally, the GO terms not common for both frameworks were mapped to zero in the x- and y-axis. (E) Top 15 GO□*biological processes* categories enriched for genes up-regulated in PD obtained with frameworks “A” and “B”, under FDR < 0.05. Bars represent log10 transformed adjusted p-values. (F) GO□*cellular component* categories enriched for genes down-regulated in PD obtained with framework “A”, under FDR < 0.1. Bars represent log10 transformed adjusted p-values. (G) Significant KEGG signaling pathways for frameworks “A” (blue) and “B” (orange), under FDR < 0.05. Framework “A” had 15 enriched KEGG pathways, from which *Chemokine signaling pathway* was also enriched in framework “B” (see Fig. S5 for the full pathway).

**Fig. 3.**
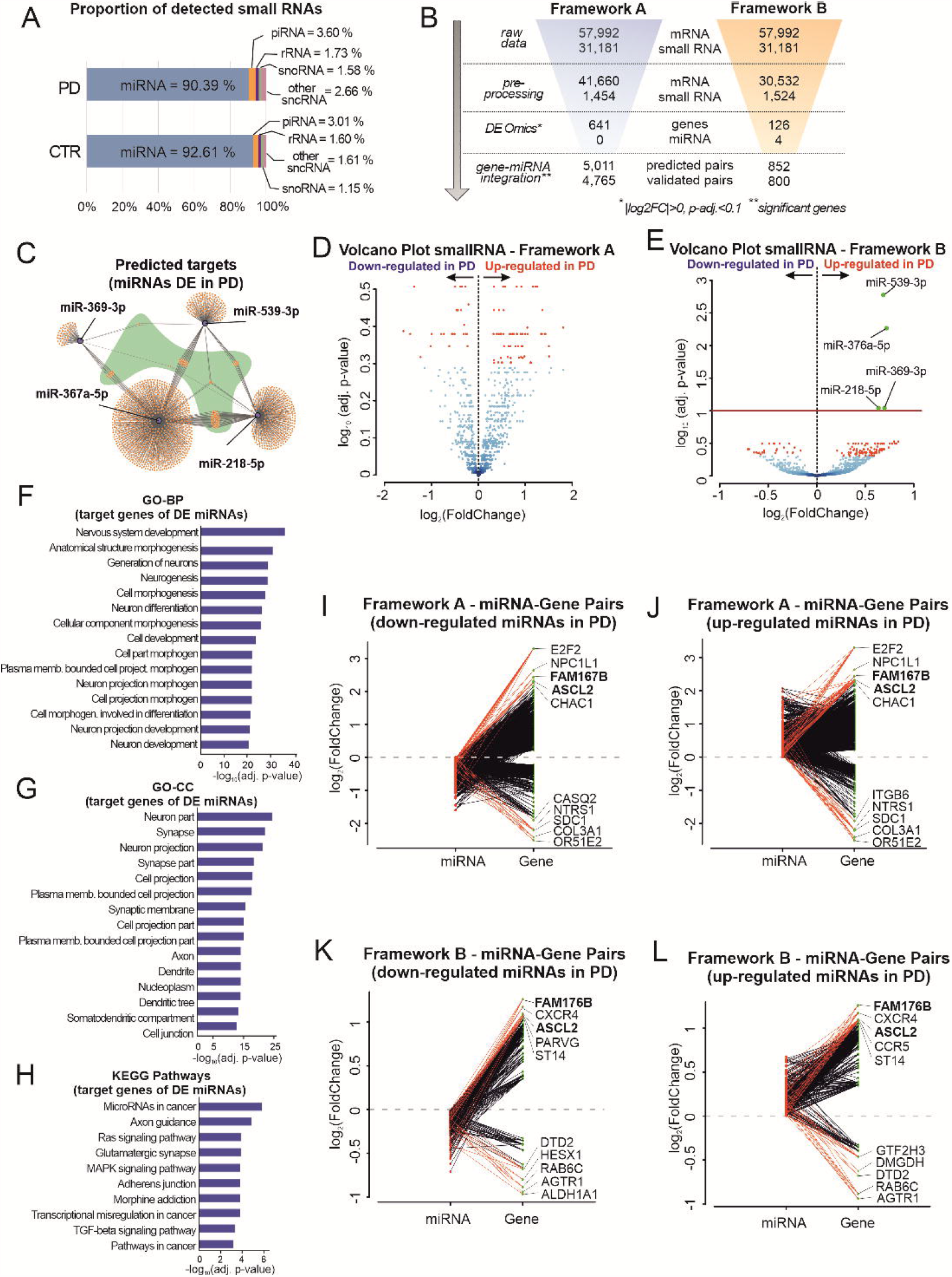
Integration of RNA sequencing experiments and analysis results of small RNA. (A) Horizontal bars depicting the percentages of the average quantities of the different small RNA species detected in the small RNA libraries as a readout for the quality of the sequencing technique for the PD patients and CTR subjects. (B) Results obtained by frameworks “A” (blue) and “B” (orange) in each step of the differential expression and integration analyses for RNA sequencing data. (C) Predicted targets for signature-miRNAs. Hub target genes that are common to the 3 miRNAs are highlighted in green. (D and E) Volcano plots portraying the differential expression of small RNA sequencing data between PD and CTR subjects, for frameworks “A” and “B”, respectively. The x-axis represents log2FoldChange(log_2_FC) and y-axis -log10(p-adjusted value). 4 up-regulated miRNAs with framework “B” were found under p-adjusted<0.1 and |log_2_FC|>0. Small RNAs attending these criteria are colored in red for positive log_2_FC. (I-J;K-L) Differentially expressed genes obtained from frameworks “A” and “B”. All mapped microRNAs (miRNAs) were integrated with their respective validated targets. For each panel, the analysis for up- and down-expressed miRNAs in PD is depicted. The y-axis denotes the log2FoldChange of the miRNAs (in red) and genes (in green). From these pairs, we highlighted genes based on their high differential level. (F,G,H) Top 15 GO□GO-BP, GO□GO-CC and *KEGG Pathway* terms enriched for the predicted targets of the differentially expressed miRNAs, respectively, under FDR<0.05. All bars represent log10 transformed adjusted p-values.

In addition, RNA sequencing data decomposition was performed (**Supplementary methods**). Data deconvolution disclosed several immune cell types that populated the analyzed samples. Interestingly, there was a decrease in *common myeloid progenitors* (log_2_FC=-49.8) with a simultaneous enrichment of *granulocyte-monocyte progenitors* in patients with PD (log_2_FC=51.6), indicating an infiltration/proliferation or increased differentiation towards the granulocyte/monocyte lineage in the analyzed midbrains (**Table S3**).

Functional annotation of the DE genes in PD revealed several biological processes that were shared between frameworks that have been known or suspected to be relevant to the pathogenesis of PD (**Fig.2D-G**). Most of these were related to *immune and inflammatory responses* (53% and 42% for each framework, respectively)(**Fig.S5**). Subsequently, terms related to *response to stress/apoptosis* and *metabolic/biosynthetic processes* characterized the DE genes obtained with framework “A” (each accounting for 5% of all enriched terms)(**Fig.S5A**), whereas *differentiation and development* and *cytoskeleton organization* were identified for framework “B” (8% and 6% of the enriched categories, respectively)(**Fig.S5B**). The pathway enrichment analysis using *Kyoto Encyclopedia of Genes and Genomes* (KEGG) included several pathways involved in infectious diseases (*HTLV-1 infection, Tuberculosis, Toxoplasmosis*), but also cancer-related processes (*transcriptional misregulation in cancer, JAK-STAT-signaling, NF-kappa B signaling)*, and several immune-related pathways (*cytokine-cytokine receptor interaction, B-cell receptor signaling pathway)* for the up-regulated genes from framework “A” (**Fig.2G**). One commonly enriched KEGG pathway, *chemokine signaling pathway*, was identified for both frameworks. For the down-regulated genes, which were considerably fewer than the up-regulated ones, no *Gene Ontology - Biological processes* (GO-BP) term reached significance (at FDR < 0.1) in any of the frameworks. Nevertheless, *Gene Ontology-Cellular compartment* (GO-CC) enrichment revealed several mitochondria-related processes for the down-regulated genes obtained with framework “A” (**Fig.2F**).

In order to explore the biological role of the four deregulated miRNAs obtained using framework “B”, we performed functional enrichment analyses with their predicted target genes (**Fig.3C**). Neuron-related pathways were the most common among enriched GO categories (**Fig.3F-H**): *neuron system development* and *neurogenesis* related pathways were the most significant GO-BP terms, while *neuron part, synapse* and *neuron projection* were the most enriched GO-CC annotations. Moreover, KEGG pathways related to *axon guidance* and *glutamatergic synapse* were also highly significant for the target genes of deregulated miRNAs. A network visualization (**Fig.S7**) disclosed eminently enriched KEGG pathways with several shared genes, e.g. *MAPK* and *Ras* signaling pathways.

For integrative assessment of the RNA sequencing datasets, we conducted a miRNA-target prediction analysis of the four significantly DE miRNAs using databases for validated and predicted targets. We identified six predicted and four validated target genes that were regulated in our total RNA sequencing data (**Table S4**). Among the identified pairs, miR-369-3p/GTF2H3 and miR-218-5p/RAB6C presented a discordant expression with an up-regulation of the miRNAs and down-regulation of transcripts in PD, representing a valid interaction between miRNAs and their target genes. For exploratory purposes, we performed an integrative analysis that considered significant genes and all of the mapped miRNAs (independent of their significance level) for both frameworks. Here, most of the regulated genes had a corresponding targeting miRNA as a valid interactor (discordant expression)(**Fig.3I-J**,**K-L)**. The expression levels of the most variant miRNAs and genes with valid miRNA pairs for both discovery and validation cohorts were visualized using boxplots (**Fig.S8**). Framework “A” yielded 4,795 unique pairs of miRNAs and gene symbols (2,459 with concordant levels of expression, and 2,336 discordant ones), whereas framework “B” yielded 800 pairs (346 with concordant levels of expression and 454 discordant ones)(**Fig.3B**,**I-J**,**K-L**).

### Protein expression profiling, functional annotation, and integration with RNA sequencing data

In order to profile the proteomics changes in our samples, we subjected midbrain tissues of our discovery cohort to Sequential Window Acquisition of all Theoretical Mass Spectra (SWATH-MS)(**Fig.S2B**). Following preparation of an annotated peptide spectral library, we were able to detect and quantitate a total of 2,257 proteins across all samples at 1% FDR.

DE analyses after normalization for the discovery cohort revealed 22 significantly regulated proteins between the PD and CTR groups. In the PD group, 17 proteins were up-regulated while five proteins were down-regulated (**Fig.4A**,**Supplementary Dataset 3**). Analysis of protein-protein interaction (PPI) networks including all significantly regulated proteins revealed several hub proteins and interactors, including TH, SELENBP1, FAH, FABP5, HSPA1B, MAP2K2, FNIP2, and PRDX1 (**Fig.4B**). The significantly regulated proteins were matched to the transcriptomics results (**Fig.4C**,**4D**), and the four most regulated genes (up- and down-regulated in PD) with their respective significant protein products were highlighted (up-regulated: CHI3L1, DNAJB1, C1QC, and SERPINA1; down-regulated: ALDH1A1, ACTA2, TAGLN, DES; (|log_2_FC|>1.4). Only Chitinase-3-like-protein-1 (CHI3L1) appeared significantly up-regulated in the PD group, in both proteomics and total RNA sequencing. Similar to the integration of transcripts and miRNA, all mapped proteins were now related to the transcriptomic analysis and miRNA data, and this revealed several links with previous datasets (**Fig.4E-H**). Functional enrichment of proteins up-regulated in PD showed the involvement of pathways related to inflammatory and immune responses (12 of the top 20 most significantly enriched categories)(**Fig.4I**). Despite the small number of down-regulated proteins in the PD group, high-level GO term grouping yielded a functional enrichment to the categories r*esponses to stress, cellular localization*, and *regulation of molecular function* (**Fig.4J**). Consecutively, the expression of selected candidates was verified in an independent cohort of six PD and two CTR subjects (**Fig.S8**,**S9**).

**Fig. 4.**
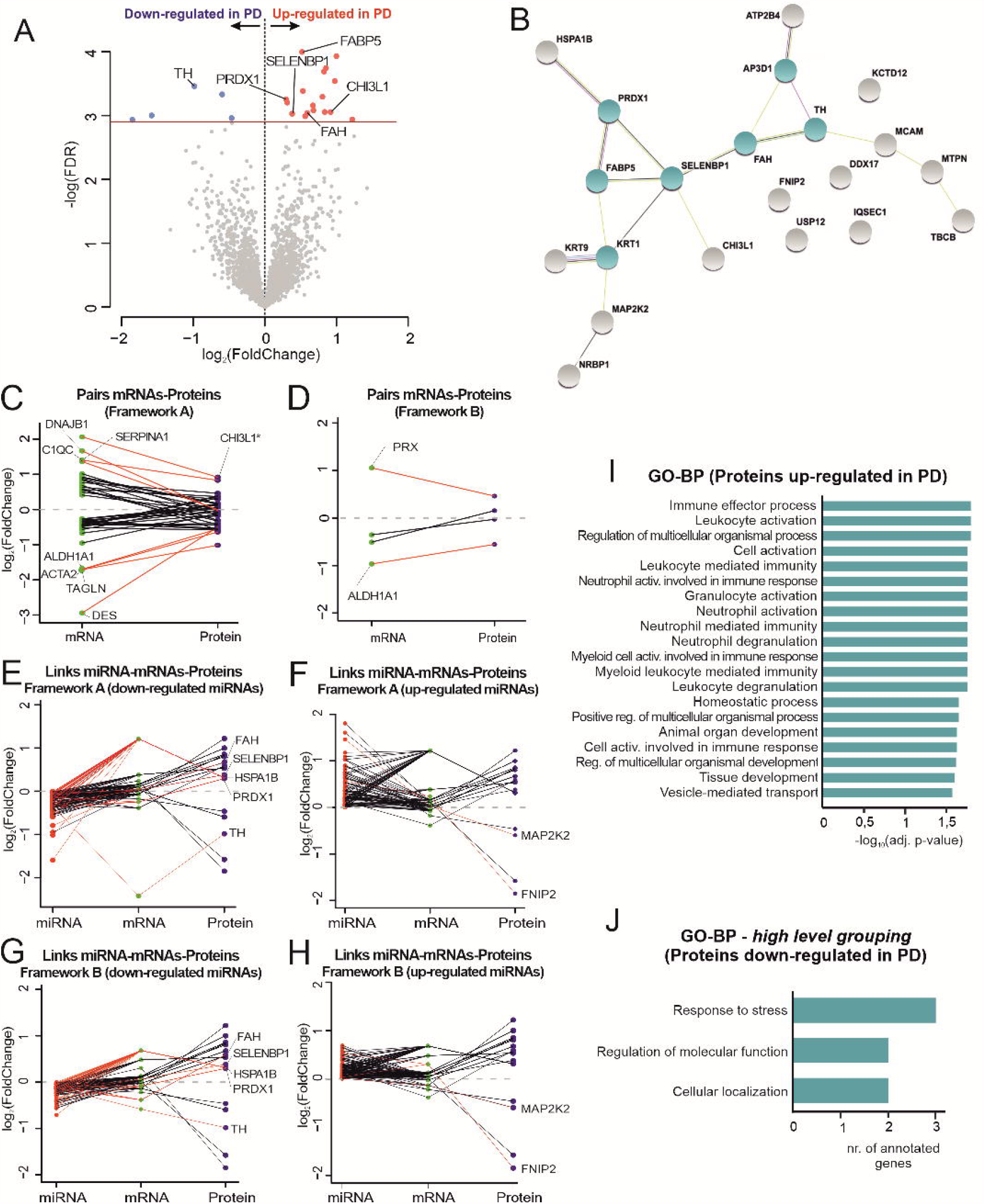
Proteomics analyses of midbrain samples. (A) Volcano plot showing all detected proteins in midbrain samples of PD and CTR subjects. Differentially expressed proteins (total of 22) between CTR and PD indicated in blue (down-regulated in PD) and in red (up-regulated in PD). Horizontal line depicts cut-off for significance (FDR=0.1). (B) STRING analysis for the differentially expressed proteins. Clusters were defined by the Markov Algorithm in STRING 11.0, using default settings. Hub proteins (3 or more links) highlighted in green. (C-D) Differentially expressed genes obtained from frameworks “A” and “B” integrated with all mapped proteins. The y-axis denotes the log2Fold Change of the genes (in green) and proteins (in purple). From these pairs, genes are highlighted based on their high differential level (top 4 genes with largest differential level, |log_2_FC|>1.4). (*)highlights CHI3L1, a candidate identified significantly up-regulated in both transcriptomics and proteomics datasets. (E;H) Combination of the resulting pairs of small RNAs and their respective target genes from frameworks “A” and “B” (independently of their significance), with the 22 significantly expressed proteins. The y-axis denotes the log2(fold change) of the miRNAs (in red), genes (in green) and proteins (in purple). (I;J) Enriched GO□BP categories for up- and down-regulated proteins in PD, respectively.

## Discussion

The lack of disease-modifying therapies in PD is a constant reminder of the need to better understand the molecular mechanisms underlying the pathology of PD in order to develop effective and etiology-driven therapies for this neurodegenerative disorder. Profiling the molecular landscape of PD-affected brains may provide insight into the pathological events taking place in the course of the disease, both at the cellular and systemic levels, and to pinpoint molecular pathways with novel druggable targets. In this study, we leveraged the potential of multi-omic tissue analysis to characterize pathology-related molecular alterations in postmortem midbrain tissue of patients with PD and age-correlated control subjects.

As our analysis aimed to identify pathways that characterize sporadic PD, we first excluded the presence of genetic alterations, that are known to cause monogenic forms of PD, in our PD cohort. Comprehensive mutational screening using gene panel sequencing and by MLPA indicated the absence of genetic abnormalities for known PD genes in the discovery cohort (**Fig.S1;Table S1**). A single-nucleotide variant of uncertain significance in the *POLG* gene was identified in one PD patient who belonged to the validation cohort; therefore, it was not included in the main analysis. Similar nucleotide variances in the *POLG* gene have been linked to alterations in PD predisposition in Finnish and Chinese populations, but mechanistic evidence of its contribution to the pathophysiology of PD is lacking (31, 32). Our findings thus confirm that the discovery cohort is composed of idiopathic PD cases and exclude a major influence due to genetic alterations in our profiling results.

Unbiased hierarchical clustering analyses of all samples demonstrated a high heterogeneity and did not permit clustering of subjects according to disease entity (**Fig.1C-H)**. In the clustering analysis of Patients with PD only, the transcriptomics analysis showed the highest homogeneity among subjects, whereas small RNAs and proteins were more heterogeneous (**Fig.1I-K**), which could argue for changes induced by post-translational modifications. The number of post-mortem samples in this project was too small to identify distinct patient subgroups; however, we recently analyzed the diversity of miRNA in cerebrospinal fluid (CSF) of patients with PD, and this revealed distinct molecular subgroups that were independent of clinical phenotype (33). A future analysis of a larger number of samples could be more suited to identify distinct subgroups and will represent an important prerequisite for the development of personalized therapeutic approaches based on the molecular phenotype in PD.

Differential expression analysis of RNA sequencing data was conducted using two different frameworks for the pre-processing stage (**Fig.2A**), with each one having a different focus. While framework “A” considered a distribution-driven approach that was appropriate for sparse data, framework “B” had a variance-based supervised procedure. A considerable number of genes were found to be significantly regulated in framework “A”, while no statistically significant miRNAs could be detected. In contrast, application of framework “B” resulted in multiple regulated genes and miRNAs (**Fig.2D**). Analyzing the RNA sequencing data using two different frameworks permitted a holistic view on the datasets, revealing subtle levels of deregulation that might not have been captured in case only one or the other was employed. Overall, this depicts the importance of using the most suitable bioinformatic approach depending on the dispersion of expression data (**Fig.S4**).

Differential transcriptome analyses revealed that there are more up-than down-regulated genes in PD (**Fig.2B-C**), which is in line with a recent meta-analysis on substantia nigra transcriptome (34). Transcriptomic overexpression was on average more common in the PD groups both at the single gene level and also when analyzing chromosomal segments with Expressed Sequencing Tags. Interestingly, despite the different transcripts obtained with each framework, both the regulation directionality and the enrichment results were very similar in both settings. Up-regulated genes in PD were enriched predominantly to inflammatory responses/immune system activation (**Fig.2E**,**Fig.S5A-B**). Processes related to *immune cell proliferation* and *defense response* were more enriched in framework “A” than they were in “B”, which might be due to the smaller number of significantly regulated genes yielded by the latter (**Fig.2D**). Similarly, KEGG pathway enrichment results are related either to inflammation/immune system activation, or to a variety of infectious diseases (**Fig.2G**), likely because pathways of infectious diseases contain genes related to immune cell activation and inflammation. One particular KEGG pathway, *chemokine signaling pathway*, which is directly related to inflammation/leukocyte recruitment (35), was shared by both frameworks (**Fig.2G**,**Fig.S5**).

Moreover, transcriptomics analysis through framework “A” also yielded functional enrichment for *stress/apoptosis* and *metabolic/biosynthetic* processes (**Fig.S5A-B**), reflecting features of neurodegeneration occurring in PD-affected midbrains (2). Metabolic dysfunction has been frequently linked to the pathogenesis of PD (36) and dopaminergic neurons are known to be especially sensitive to oxidative stress/mitochondrial dysfunction (37). Strikingly, PD down-regulated genes also unveiled an enrichment in numerous mitochondrial processes, which are known to be markedly impaired in autosomal recessive forms of PD (38)(**Fig.2F**). Framework “B” led to enrichment for the category *differentiation and development*, and a similar enrichment was observed for the deregulated miRNAs. Cell cycle-related proteins play an important role in the survival of mature neurons, and also in neuronal apoptotic processes (39). Overall, those findings support the connection between the miRNA and gene expression datasets, encouraging the integrative efforts employed in this study.

For the small RNA data, we were able to identify 4 differentially expressed miRNAs using framework “B”, all of which were up-regulated in PD (**Fig.3G**). Similar findings also suggested an overall up-regulation of circulating miRNAs in PD (40, 41). Remarkably, all candidates depicted here have already been linked to PD and other neurodegenerative diseases. miR-218-5p was found be up-regulated in peripheral blood mononuclear cells (PBMCs) of patients with PD (42). Its levels were reversed after deep brain stimulation (DBS), suggesting that DBS therapy directly influenced miR-218-5p expression. miR-218-5p was also found to be altered in the CSF of Alzheimer’s disease (AD) patients, but in decreased levels (43). Furthermore, miR-376a-5p was found in increased levels both in PD PBMCs and *in vitro*. Interestingly, it regulates genes that are involved in mitochondrial dysfunction and were previously linked to PD pathogenesis (TFAM; PGC1α)(44). miR-376a-5p also regulates SIRT2 levels (45), a protein implicated in PD for its effects in α-syn aggregation (46). It has also been postulated as a biomarker for PD, since its levels in PD PBMCs also seem to correlate with disease severity (44). One study showed elevated levels of miR-369-3p in PD postmortem substantia nigra (47), while alterations in the striatal levels of miR-539-3p were reported in a rodent model of PD (48). Functionally, these miRNAs were majorly linked to neuron-related gene ontology terms, indicating the neuronal origin of these miRNAs (**Fig.3J-L**). Additionally, terms related to cell-cycle, proliferation, differentiation, and development were greatly enriched. These are known to also play a vital role in the survival and maintenance of mature neurons (49) and are directly influenced by miRNA levels during neurodegeneration (50). Aberrant cell-cycle was also reported to be intimately linked to neurodegeneration (51). Generally, functional enrichment for deregulated miRNAs indicate that the pathological pathways captured here are intimately related to neuronal insults in the context of PD.

To obtain a more holistic view, the RNA sequencing datasets comprising transcriptome and microRNAome were analyzed in an integrative fashion (**Fig.3E-I**). PD up-regulated genes included transcription factor E2F2, implicated in hemisphere-dependent PD pathology (52) and neuroinflammation after spinal cord injury (53); Glutathione-specific gamma-glutamylcyclotransferase-1 (CHAC1), reported to regulate unfolded protein response (54) and linked to paraquat-induced neurotoxicity (55); and CXCR4, a chemokine receptor intimately linked to neuroinflammation which is expressed in dopaminergic neurons (56). DRD2 and OR51E2 presented marked down-regulation in PD. The former has been found in altered levels in PD substantia nigra and has been regarded as a susceptibility locus for PD (34, 57), whereas the latter is postulated as a therapeutic target for PD. It is involved in neuromelanin pigmentation in dopaminergic neurons and was found to be down-regulated in PD-affected brains (24, 58).

When considering only significantly differentially expressed miRNAs and transcripts, the list was reduced to two valid miRNA-target gene pairs: miR-218-5p/RAB6C and miR-369-3p/GTF2H3 (**Table S4**). The ras-related protein Rab-6C (RAB6C) is a member of Rab GTPases, which are pivotal regulators of intracellular protein transport and vesicle trafficking (59). RAB6C has been implicated in relevant processes associated with PD, including autophagy and proteostasis. Increased levels of Rab6 isoforms have been linked to protein aggregation *in vitro* (60), and RAB6C is postulated to be a modulator of the unfolded protein response in AD-affected brains (61). Furthermore, a recent study has shown that Rab GTPases are substrates of LRRK2, the most commonly mutated gene in familial cases of PD. Disruption of Rab phosphorylation in the LRRK2 site leads to neurotoxicity *in vitro* and dopaminergic neurodegeneration *in vitro* (62). The general transcription factor IIH subunit 3 (GTF2H3) is a transcription factor that is involved in RNA transcription and nucleotide excision repair (63). Although GTF2H3 has not been directly implicated in PD, dysfunctions in nucleotide excision repair play an important role in chronic neurodegenerative disorders (64). Moreover, members of its family (TFIIH) have been shown to play a role in oxidative stress and mitochondrial DNA alterations in PD (65). Altogether, these candidates underline pathways that are involved in the pathogenesis of PD and support the involvement of gene expression regulation by miRNAs as a disease mechanism. Furthermore, the regulation of these targets is supported by the boxplots built for the validation cohort (**Fig.S8**). Finally, proteomic profiling ensures a better characterization of disease-relevant mechanisms taking place post-translationally. In line with the transcriptomic data, most proteins were up-regulated in PD (**Fig.4A**). As expected, differential expression results revealed a down-regulation in tyrosine hydroxylase (TH), the rate-limiting enzyme for dopamine synthesis and a hallmark of dopaminergic neuron depletion in PD (2, 66). PPI networks revealed other important hub-proteins (**Fig.4B**), such as the selenium-binding protein 1 (SELENBP1), fumarylacetoacetase (FAH), fatty acid-binding protein 5 (FABP5), and peroxiredoxin-1 (PRDX1). Remarkably, we previously identified selenium as part of a bioelement-signature in the CSF that allowed for differentiation between PD and control samples (67). FAH and PRDX1 were reported to play a role in oxidative stress damage in PD, and the latter has also been postulated as a new therapeutic target for PD because of its relationship with the generation of reactive oxygen species (68, 69). FABP5 has been linked to postnatal neurogenesis (70), and also to neuronal oxidative damage in vitro (23). Another important protein that was found to be up-regulated in PD was heat shock 70 kDa protein 1B (HSPA1B), a chaperone known for its role in protein folding and degradation as well as neuronal apoptosis (71). Chaperones are known to interact with αSyn (72), and defects in that system contribute directly to αSyn misfolding and the formation of inclusions/protein aggregates in PD-affected brains (73). Significantly regulated proteins were substantially fewer than deregulated transcripts, most likely because only the most abundant/uniquely identified proteins within all samples were considered for quantification. Furthermore, proteomics and RNA sequencing results are fundamentally very different, starting with the nature of the signals and the coverage of those techniques, which often limits the overlap between such datasets (74). Nevertheless, their integration resulted in a better understanding of the regulation of identified candidate molecules, for example by comparison of individual protein expression levels to respective RNA sequencing results (**Fig.4C-D**) and by analysis of regulation in the validation cohort (**Fig.S8**).

An important finding was the identification of chitinase-3-like-protein 1 (CHI3L1) which was up-regulated in both transcriptomics and proteomics datasets. It showed the highest levels of up-regulation in both analyses after integration (**Fig.4C**). CHI3L1, also known as YKL-40, is widely expressed in immune cells and plays a major role in the regulation of inflammatory responses, tissue injury and repair (75). Remarkably, it has been previously identified in CSF studies as a potential circulating biomarker for several neurodegenerative diseases, including Alzheimer’s disease (AD), amyotrophic lateral sclerosis (ALS), and PD (76–78). It was also considered as a pivotal marker for immune/inflammatory changes in tauopathies (79). Functional analysis revealed the GO-BP *immune effector process* as the most enriched term for the up-regulated proteins in PD (**Fig.4I**), matching the massive immune/inflammatory activation reported for RNA sequencing results and the deregulation of several immune mediators in the proteomics dataset. Remarkably, functional alterations related to neuroinflammation/immune response activation were captured across all omics results presented in our study and were independent of data handling paradigms. Neuroinflammation has already been considered to be a predictive feature for the appearance of non-motor symptoms and cognitive decline in patients with PD (80), and inflammation-related mechanisms seem to accompany the pathophysiological events of PD even from earlier stages (5, 9). A recent study showed that neuroinflammatory mechanisms are triggered with the release of αSyn from apoptotic neurons, aggravating the disease through a number of pathways that include microglial activation, mitochondrial damage and inflammasome formation (81). Our results provide further evidence for the role of neuroinflammation in PD pathology. However, we must caveat that our data are representative of only advanced stages of PD, since the analyzed postmortem samples were obtained from patients with disease duration of 7–25 years.

It is important to highlight that factors modulating protein metabolism (e.g., synthesis and degradation rates of proteins) were out of the scope of this study and were therefore not considered for the integrative analyses presented here, although they are likely to significantly contribute to the overall picture. The analysis of the phospho-proteome and the metabolome was also not included because of the postmortem nature of the source material. Limitations also apply to the number of analyzed subjects, because high-quality post-mortem tissues with excellent clinical characterization are scarce, particularly for control subjects without neurodegenerative pathology. However, as all analyses showed a strong focus on neuroinflammation, we do not expect that the inclusion of additional samples will change this overall picture. Nevertheless, an analysis of larger cohorts will be much more powerful for subgroup identification and may identify additional molecular targets characterizing subtypes of PD pathology.

In summary, our integrative multi-omics analyses identified multiple levels of deregulation in human midbrains affected by PD, with several of them overlapping across the different generated datasets. The overwhelming representation of neuroinflammation-related molecules suggests a pivotal role for neuroinflammation in the pathogenesis of PD and also delineates a yet unaddressed drug target in PD (82). Moreover, we identified four deregulated miRNAs, namely, miR-539-3p, miR-376a-5p, miR-218-5p, and miR-369-3p that have been previously linked to neurodegenerative processes, providing additional evidence of the importance of these candidates in the context of PD pathology. We also found a number of miRNA-target gene interacting pairs, including the valid pairs of miR-218-5p/RAB6C and miR-369-3p/GTF2H3, that underline the contribution of vesicle trafficking, proteostasis, and oxidative stress in the pathogenesis of PD. Canonical PD-related proteins, such as TH, were also captured in our study, alongside further protein candidates, such as CHI3L1, SELENBP1, PRDX1, and HSPA1B. Further mechanistic and clinical studies would be important to substantiate the pathological importance of our findings in the context of earlier disease stages.

## Materials and Methods

### Human postmortem midbrain samples

Human midbrain samples were provided by the Parkinson’s UK Brain Bank (Imperial College London, London, England). In total, 19 PD and 12 CTR samples were obtained, shipped, and processed in two different batches. Frozen midbrain tissue blocks were transported and stored under controlled temperature conditions. The samples were conceded to the Department of Neurology, University Medical Center Göttingen, Göttingen, Germany, and ethical approval was given by the Multicenter Research Ethics Committee (07/MRE09/72). **Table 1** encloses all clinical information about the subjects. For sampling, midbrain blocks were transferred to a cryostat chamber and kept at -20°C. Each block was punched with a 20-G Quincke Spinal Needle (Becton Dickinson), ∼20 mg tissue was collected into reaction tubes and kept at -80°C until further use.

**Table 1.** Demographic and clinical characteristics of analyzed patient cohorts. Data are presented as mean±SEM, and values in squared brackets represent the range. Statistical tests for nominal variables (sex) were performed using Fisher’s exact test and for continuous variables (age and PMI) leveraged Student’s t-test, under a significance level of 5%. Differences between CTR and PD groups within each cohort not significant for sex (p-value=0.4136 and p-value=0.4286, for the discovery and validation cohorts, respectively) and age at death (p-value=0.7893 and p-value=0.6731, for the discovery and validation cohorts, respectively). PMI values were significantly different in the discovery cohort (p-value=0.00811), but not in the validation cohort (p-value=0.07209). These differences were not reflected in RNA library size. PMI:postmortem interval.

|  | discovery cohort |  | validation cohort |  |
| --- | --- | --- | --- | --- |
|  | CTR | PD | CTR | PD |
| number of patients | 10 | 13 | 2 | 6 |
| sex (m/f) | 4/6 | 8/5 | 0/2 | 4/2 |
| age at death | 77.1 ± 3.4<br>[61–96] | 78.3 ± 1.3 [71–87] | 74.5 ± 5.3<br>[60–89] | 82.7 ± 1.7<br>[73–93] |
| disease duration (years) | NA | 13.4 ± 1.3<br>[7–25] | NA | 12.2 ± 1<br>[7–19] |
| postmortem interval (PMI) | 20.6 ± 1.2<br>[12–25] | 14 ± 1.4<br>[3–24] | 13<br>[13] | 17.5 ± 1.1<br>[10–24] |

### RNA and DNA isolation from human midbrain samples

Total RNA was isolated from human using TRIzol (Invitrogen) following the manufacturer’s instructions (**Supplementary methods**). After extraction, RNA samples were incubated at 55°C for 2 min in order to completely dissolve the RNA, and DNAse treatment (Life Technologies) was performed. RNA samples were cleaned with the RNA Clean&Concentrator-5 Kit (Zymo Research). RNA integrity was assessed with the Agilent 6000 NanoKit in the 2100 Bioanalyzer (Agilent). DNA isolation from human midbrain samples was performed with the QIAamp DNA Mini Kit following the manufacturer’s instructions.

### RNA sequencing experiments

RNA sequencing was performed in the NIG-NGS Integrative Genomics Core Unit, University Medical Center Göttingen. Small RNA libraries were prepared using the TruSeq Small RNA LibraryPrep Kit (Illumina) with minor modifications (**Supplementary methods**). The quality/integrity of RNA libraries was assessed in the Fragment Analyzer (Agilent). All sequenced samples exhibited comparable RNA integrity. Both small and total RNA sequencing were performed on the Illumina HiSeq4000 platform (Illumina), generating 50bp single-end reads (small RNA sequencing: 10–20 million reads/sample; total RNA sequencing: 30–40 million reads/sample). After sequencing, sequence images were transformed to BCL files with the BaseCaller software. The files were demultiplexed to fastq files with bcl2fastq v2.17.1.14.(Illumina). Quality check was done with FastQC v.0.11.5.

### RNA sequencing data processing and mapping

After small and total RNA sequencing, the data was processed with a customized in-house pipeline (**Supplementary methods**). After adapter trimming/demultiplexing, reads were mapped to the reference genome (miRNAs/piRNAs/other non-coding RNAs known sequences). The reads were mapped in the non-splice-junction-aware mode. No mismatches for the reads <19b were allowed. For reads between 20b-39b, one mismatch was allowed, and for reads between 40b-59b, two mismatches were tolerated. All other parameters were set as default in RNA-STAR.

### Sequential window acquisition of all theoretical mass spectra (SWATH-MS)

After protein lysate preparation with Urea/Thiourea/Chaps lysis buffer (details in **Supplementary methods**), 50µg protein were loaded into a 4–12% NuPAGE Novex Bis-Tris Minigels (Invitrogen). Following electrophoresis, the bands stained with Coomasie Brilliant Blue (ThermoFisher) were cut-out, diced, added of dithiothreitol alkylated with iodoacetamide for reduction, and digested with trypsin overnight. Tryptic peptides were extracted from the gel and the solution was dried in a Speedvac. After spectral library generation (**Supplementary methods**), protein digests were analyzed on an Eksigent nanoLC425nanoflow chromatography system (AB Sciex) hyphenated to a hybrid triple quadrupole-TOF mass spectrometer (TripleTOF 5600+). Qualitative liquid chromatography/tandem mass spectrometry (LC-MS/MS) analysis was performed using a Top25 data-dependent acquisition method (**Supplementary methods**). Three technical replicates per reversed-phase fraction were analyzed to construct a spectral library. During quantitative SWATH analysis, three replicate injections were acquired for each sample.

### Mass spectrometry data processing

Protein identification was achieved using Protein Pilot Software v.5.0 build4769 (AB Sciex) at thorough settings. Spectral library generation and SWATH peak extraction were achieved in PeakView Software version 2.1 build 11041 (AB Sciex) using the SWATH quantitation microApp (v.2.0 build2003). Following retention time correction by the iRT standard, peak areas were extracted using information from the MS/MS library at a false discovery rate (FDR) of 1% (83). Finally, the resulting peak areas were summed to peptide area values and next to protein area values.

### Differential expression analyses of small and total RNA sequencing data

Two complementary computational frameworks based on DESeq2(84) were used for the differential expression analysis of small/total RNA-seq data. Pipelines differed in the pre-processing procedure (**Supplementary Methods**). Adjusted p-value of<0.1 / absolute log_2_FoldChange (|log_2_FC|)>0 were considered as significance cut-offs. The Grubb’s-test (at 0.05 significance level) was used to identify putative outlier samples.

### Proteomics differential expression analyses

The differential expression analysis of the proteomics was performed with Perseus(85). This application performs multiple hypothesis testing correction using a permutation-based False Discovery Rate (FDR) approach, where Analysis of Variance (ANOVA) and p-values are computed between the measured and permuted data using Student’s t-test statistical hypothesis test. FDR values were calculated as fractions of accepted hits from the permuted data over the measured one. Proteins were considered statistically differentially expressed with an FDR<0.1.

### MiRNA target prediction and multi-omics data integration

After target mining, miRNAs identified by small RNA sequencing were matched to target transcripts identified in the parallel total RNA sequencing experiments. Details about miRNA-target prediction depicted in **Supplementary methods**. Similarly, integration between gene and protein expression data was done for genes identified in the total RNA sequencing experiments with a protein product also identified by SWATH-MS experiments.

### Gene ontology and pathway enrichment analyses

GO term and pathway enrichment analyses were performed using and several common functional annotation databases (i.e., GO, KEGG, STRING). Details about selected databases and scripts depicted in **Supplementary methods**. Enrichment analysis was done separately for up-/down-regulated entities under a significance level of FDR<0.05 and <0.1, respectively.

## Data Availability

RNA sequencing and proteomics datasets were deposited in public repositories (the European Genome-phenome Archive (EGA) and the Proteomics Identifications Database (PRIDE), respectively) following applicable guidelines. RNA sequencing data are stored under the accession number EGAD00001006883. Proteomics data are available via ProteomeXchange with the identifier PXD023618.

## Declarations

### Ethics approval-sample acquirement

All human midbrain samples were provided by the Parkinson’s UK Brain Bank (Imperial College London, London, England). The samples were conceded to the Lingor Lab (Department of Neurology, University Medical Center Göttingen, Göttingen, Germany). Ethical approval was given by the Multicenter Research Ethics Committee (07/MRE09/72).

### Data availability

RNA sequencing and proteomics datasets were deposited in public repositories—the European Genome-phenome Archive (EGA) and the Proteomics Identifications Database (PRIDE), respectively—following applicable guidelines. RNA sequencing data are stored under the accession number EGAD00001006883. Proteomics data are available via ProteomeXchange with the identifier PXD023618.

### Code Availability

All bioinformatics analyses were performed using R software v.3.6.0 (R Foundation for Statistical Computing, Vienna, Austria). The source code is available at https://github.com/AnaGalhoz37/Multi-Omics-PD.

## Acknowledgments

The authors thank the Parkinson’s UK Brain Bank for conceding the postmortem midbrain samples for this study, as well as the NIG-NGS Integrative Genomics Core Unit, University Medical Center Göttingen for performing RNA sequencing experiments, and Centogene AG for conducting the gene panel analyses.

## Figure legends – Abbreviations

CTR: control
PD: Parkinson’s disease
PMI: postmortem interval
Age: age at death
NP diagnosis: neuropathological diagnosis
Gender: F-females/M-males
LBDBS: Lewy body disease brainstem variant
LBDE: Lewy body disease early-neocortical stage
LBDN: Lewy body disease neocortical stage
miRNA: microRNA
GO: Gene Ontology
BP: biological process
CC: cellular compartment
KEGG: Kyoto Encyclopedia of Genes and Genomes
FDR: false discovery rate
DE: differentially expressed
FC: fold change
piRNA: Piwi-interacting RNA
rRNA: ribosomal RNA
snoRNA: small nucleolar RNA
sncRNA: small non-coding RNA

