## Supplemental Information for "Multi-omic landscaping of human midbrains identifies neuroinflammation as major disease mechanism in advanced-stage Parkinson’s disease"

**Supplementary Information Text**

**Supplementary Methods**

**DNA Isolation**

DNA isolation from human midbrain samples was performed with the QIAamp DNA Mini Kit following the manufacturer’s instructions. Nucleic acid concentration and purity were measured in the NanoDrop One spectrophotometer (ThermoFisher, Waltham, MA, USA).

**Multiplex Ligation-dependent Probe Amplification (MLPA)**

MLPA experiments were performed in order to detect abnormalities in copy numbers (e.g. deletions, duplications, triplications) of specific PD-related genes using a set of standard commercial probes (SALSA P051 and P052 Parkinson MLPA kits, MRC Holland, Amsterdam, The Netherlands). MLPA experiments were conducted according to the manufacturer's protocol. PCR amplification products were visualized on capillary sequencing machines (either ABI 3130XL or 3500XL) (Applied Biosystems, Foster City, CA, USA) using the coffalyser software (MRC Holland, Amsterdam, The Netherlands).

**Gene Panel sequencing**

For Gene Panel analysis, DNA samples were sequenced on a next-generation sequencing platform with a collaboration partner (Centogene AG, Rostock, Germany). 29 genes previously linked to PD or dystonia (DYT) phenotypes were analyzed. The mean sequencing depth was 600x. Variants were filtered according to quality scores (cut-offs: quality score >200; coverage >10, allele fraction >20%), further filtered for the number of times that they appeared in public databases (cut-off: <0.01 in public databases and the in-house database) and finally for protein-changing variants in PD genes. Candidate variants were confirmed by Sanger sequencing. All genes included in the gene panel are disclosed in **Table S2**.

**RNA and DNA isolation from human midbrain samples**

Total RNA was isolated from human using TRIzol (Invitrogen) following the manufacturer’s instructions. 1 ml of TRIzol was added to each midbrain sample, followed by 100 µL of 1-Bromo-3-Chlor-Propane (Sigma Aldrich). Samples were mixed by inversion and the lysates were centrifuged at 12.000 × g for 15 min at 4°C, for organic/aqueous phase separation. The RNA-containing aqueous phase was isolated and RNA precipitation was performed with 500 µl of 2-propanol (AppliChem) and 2 µl GlycoBlue Co-precipitant (15 mg/ml)(ThermoFisher). Next, the samples were centrifuged at 12.000 × g for 30 min at 4°C. RNA pellets were washed three times with 75% ice-cold ethanol (AppliChem). The pellets were dried for 5 min, and reconstituted with 15-20 µl of nuclease-free water (Sigma Aldrich). RNA samples were incubated at 55°C for 2 min in order to completely dissolve the RNA. Finally, DNAse treatment (Life Technologies) was performed in accordance with the manufacturer’s instructions, and the RNA samples were cleaned and concentrated with the RNA Clean & Concentrator-5 KIT (Zymo Research). RNA integrity was assessed with the Agilent 6000 Nano Kit in the 2100 Bioanalyzer (Agilent). DNA isolation from human midbrain samples was performed with the QIAamp DNA Mini Kit following the manufacturer’s instructions.

**Preparation of RNA Sequencing Libraries**

The same RNA source was used for both small and total RNA sequencing experiments. Small RNA libraries were prepared using the TruSeq SmallRNA Library Prep Kit (Illumina) with minor modifications: 600 ng total RNA were used as starting material for library preparation. In order to prevent the formation of adapter dimers (by 5΄ and 3΄ self-ligation) and consequent amplification of these dimers, the CleanTag Library Preparation for Next-Generation Sequencing Kit (TriLink, San Diego, CA, USA) was employed. Total RNA libraries were prepared using a modified version of the TruSeq Stranded Total RNA protocol (Illumina), a strand-specific, massive-parallel cDNA sequencing protocol. 200 ng of total RNA were used as a start material. A ribosomal RNA (rRNA) depletion protocol (RiboMinus™, ThermoFisher, Waltham, MA, USA) was performed in order to maintain rRNA content under 5% in the samples. Next, an adaptor ligation step is performed, followed by PCR amplification of the reads. A reduced number of PCR cycles was employed in order to avoid PCR duplication artifacts, as well as primer dimers in the final libraries. The standard sensitivity RNA Analysis Kit was used for Fragment Analyzer runs. For accurate quantitation of small RNA libraries, a library pool was quantified with the QuantiFluor dsDNA System (Promega). cDNA library sizes were determined with the dsDNA 905 Kit (Agilent).

**RNA sequencing data processing and mapping**

After small and total RNA sequencing, the data was processed with a customized in-house pipeline (1). Adapter trimming and demultiplexing were then performed alongside base calling. For the small RNA sequencing data, the 3' adapters were trimmed and reads with the minimum length of 16 nucleotides were filtered out with the Cutadapt pipeline (2). The reads were then mapped to the reference genome for miRNAs/piRNAs known sequences, followed by other small non-coding RNAs. The remaining unmapped reads were mapped to the human genome. Bowtie version 1.1.2 (3) was employed for all mapping steps, and no mismatches were allowed for reads ≤ 32 b. For reads between 33 b and 50 b, one mismatch was tolerated. For total RNA sequencing data, RNA reads were mapped to the human transcriptome using RNA-STAR version STAR_2.5.2b (4) for all mapping steps. The reads were mapped in the non-splice-junction-aware mode and no mismatches for the reads <19 b were allowed. For reads between 20b and 39b, one mismatch was allowed, and for reads between 40b and 59b, two mismatches were tolerated. Besides the parameters depicted here, all other parameters were set as default in RNA-STAR. Aligned reads overlapping exons (for each gene) was counted with the intersection-non-empty mode of the htseq-count script (HTSeq package version 0.9.1)(5).

**Preparation of peptide libraries, SWATH-MS Run Settings and data mapping**

For proteomics experiments, human midbrain tissue lysis was performed with Urea/Thiourea/Chaps lysis buffer. Samples were homogenized and sonicated for 30s intervals and amplitude of 40%. Protein quantification was performed with the modified-Bradford Roti-Nanoquant Kit (Carl-Roth). For the generation of peptide libraries, equal aliquots from each sample were pooled to a total of 80 µg and further separated into eight fractions using a reversed-phase spin column (Pierce High pH Reversed-Phase Peptide Fractionation Kit) (ThermoFisher). Spike-ins from a synthetic peptide standard were added to the samples and used for retention time alignment (iRT Standard). Protein digests were analysed on the Eksigent nanoLC425 nanoﬂow chromatography system (AB Sciex), hyphenated to a hybrid triple quadrupole-TOF mass spectrometer (TripleTOF 5600+) equipped with a Nanospray III ion source (Ionspray Voltage 2400 V, Interface Heater Temperature 150°C, Sheath Gas Setting 12) and controlled by Analyst TF 1.7.1 software build 1163 (AB Sciex). Peptides were dissolved in a loading buffer (composed of 2% acetonitrile and 0.1% formic acid in water). Following column enrichment, separation was performed on an analytical RP-C18 column of dimensions 0.075 mm ID x 250 mm, HSS T3, 1.8 µm (Waters, Milford, MA, USA) using a 90 min linear gradient of 5-35 % acetonitrile / 0.1% formic acid (v:v) at 300 nl min-1. Qualitative liquid chromatography/tandem mass spectrometry (LC-MS/MS) analysis was performed using a Top25 data-dependent acquisition method. For that, an MS survey scan of m/z 350–1250 accumulated for 350 ms at a resolution of 30,000 full width at half maximum (FWHM) was performed. MS/MS scans of m/z 180–1600 were accumulated for 100 ms at a resolution of 17,500 FWHM and a precursor isolation width of 0.7 FWHM, resulting in a total cycle time of 2.9 s. Precursors above a threshold of 125 cps for MS intensity and with charge states of 2+ / 3+ / 4+ were selected for MS/MS. Dynamic exclusion time was set to 30 s. An MS/MS activation was achieved by CID using nitrogen as a collision gas. For quantitative sequential window acquisition of all theoretical fragment ion spectra (SWATH) analysis, MS/MS data were acquired using 65 variable size windows across the 400-1,050 m/z range. Fragments were produced using rolling collision energy settings for charge state 2+, and fragments acquired over an m/z range of 350–1400 for 40 ms per segment. With the inclusion of a 100 ms survey scan, the overall cycle time was of 2.75s. A total of 407,752 MS/MS spectra from the combined qualitative analyses were searched against the UniProtKB human reference proteome (revision 04/2018, 93.609 entries) augmented with a set of 52 known common laboratory contaminants.

**Unsupervised visualizations**

The top 100 most variant omics were identified using the rowVars function available in the matrixStats package in R software (6), followed by a z-score transformation. Subsequently, we performed a complete hierarchical clustering over all samples, and resulting heatmaps were illustrated using the ComplexHeatmap visualization package (7). Furthermore, pairwise sample correlation analyses for small and total RNA sequencing, and proteomics expression were performed using Bayesian Hierarchical Clustering (8). For this, the PD samples of the discovery cohort and normalized mapped counts without any additional transformation were used as input (8, 9).

**Pre-processing of small and total RNA sequencing data**

Two distinct pre-processing procedures to remove low-count and low-variance data were considered. Framework A starts with an initial elimination of all zero-count-row entrances of the count matrix, and is followed by the removal of low-count small RNA-seq/genes based on their data distribution using a quantile filtering with a cutoff of 25% and 50%, respectively for the small RNA-seq and total RNA-seq data, using the TCGAanalyze Filtering function available in the R package TCGAbiolinks (10). In contrast, framework B initializes by estimating the unwanted variance in the given matrix count of genes/small RNA-seq and proceeded identification and removal of house-keeping-genes, i.e. a set of genes whose expression is not influenced by the conditions of the study, using the Remove Unwanted Variance (RUV) available in the R package RUVSeq (11). Further, the raw counts are transformed into log-form counts by assuming a Negative-Binomial (NB) distribution using the Variance Stabilizing Transformation (VST)(12) method for data visualization.

**RNA-seq decomposition**

In an attempt of making an assessment on the proportion of immune cells present in the different cohorts (PD patients and control), we leveraged the RNASeq Decomposition computational tool presented by Finotello and Trajanoski (13). For this, we computed the TPM-transformed values of our original RNA-seq count data considering a gene length of 50 bp (see library preparation section) and leveraging the R function counts_to_tpm developed by Kamil Slowikowski (14). Considering we are interested in the comparison between cohorts, we applied the deconvolution methods *xcell* and *mcp-counter* since these are based on marker genes and therefore suitable to applications on data with different conditions. Further, mean values of the scores were calculated for each cell, and subsequent log_2_FoldChange (log_2_FC) between PD and control patients scores. Significance was introduced by a Wilcoxon t-test and succeeding computed adjusted p-values based on Bonferroni’s correction method. Significant cell scores were identified by adjusted p-value < 0.1.

**MiRNA target prediction and multi-omics data integration**

To predict miRNAs target genes, we used miRDB v22 (15) and mirTarBase 8.0 (16). The first is based on text mining of functional studies of miRNAs and collects miRNA-target interactions (MTI) which have been experimentally validated. The latter consists in the application of support vector machines in high-throughput sequencing experiments, and the identification of new miRNA target binding and expression downregulation. For miRDB, the default search criteria of target prediction score higher than 60 and miRNAs with less than 800 targets was considered to find all gene targets of all the miRNAs available in our data. In addition, integration between gene and protein expression was performed using the UniProt database (17).

**Gene ontology and pathway enrichment analyses**

To perform a GO term and pathway enrichment analyses of the differential expressed genes, we used ShinyGO (18) and several common functional annotation databases (i.e., GO and KEGG). This enrichment analysis was done separately for the differentially up- and down-regulated entities, and using a significance level of FDR < 0.05 and < 0.1, respectively. Furthermore, protein-protein interaction networks were generated and visualized with the STRING platform v.11 (19) using default settings.

**Supplementary Figures:**

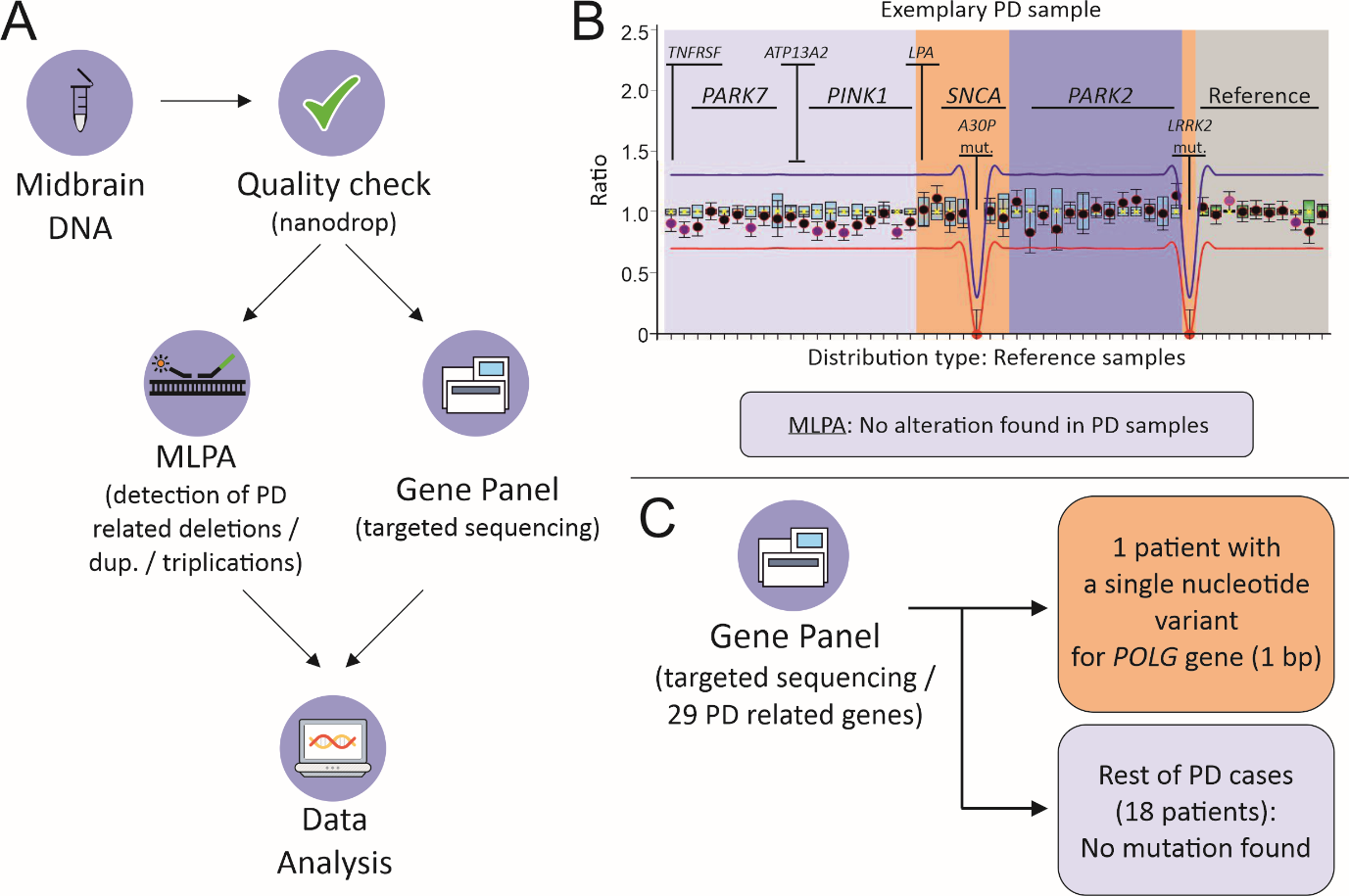

**Figure S1. Gene panel and MLPA experiments overview. (A) Experimental design. After quality check, DNA samples were processed in MLPA / Gene panel sequencing experiments. (B) Exemplary MLPA results for a PD patient showing no alteration (deletion, duplications or triplications) for the explored genes. Likewise, no alterations were found in the PD patient cohort. (C) Gene panel experiments reveal a single nucleotide variant (length: 1 bp) for the *POLG* gene in one of the PD patients. No mutations found for the remaining cohort.**

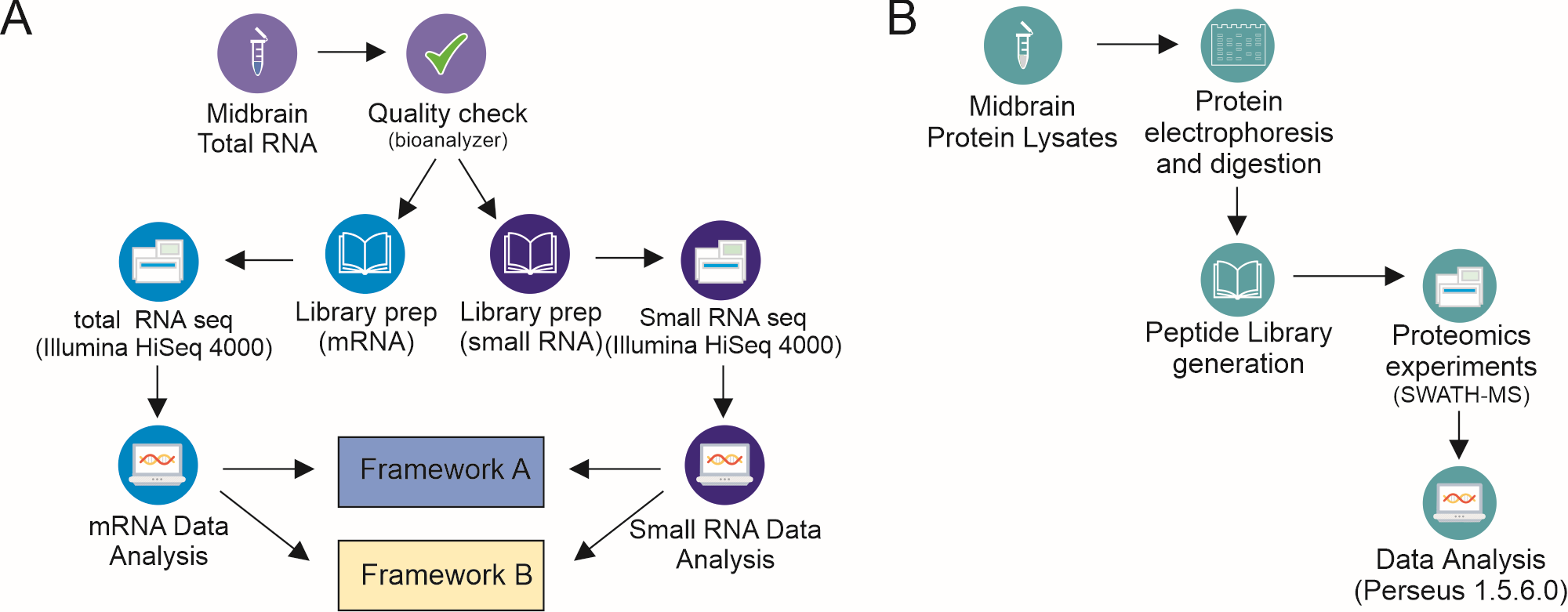

**Figure S2. Experimental design for RNA sequencing and proteomics experiments. (A) After quality check, small and total RNA libraries were prepared from each midbrain sample. Sequencing was performed in the Illumina HiSeq 400 platform, followed by bioinformatic analysis with two different frameworks (“A” and “B”). (B) Proteins extracted from midbrain tissue were separated by electrophoresis and digested by trypsinization. Peptide libraries were generated and Sequential Window Acquisition of all Theoretical Mass Spectra (SWATH-MS) experiments were performed, followed by data analysis using Perseus 1.5.6.0.**

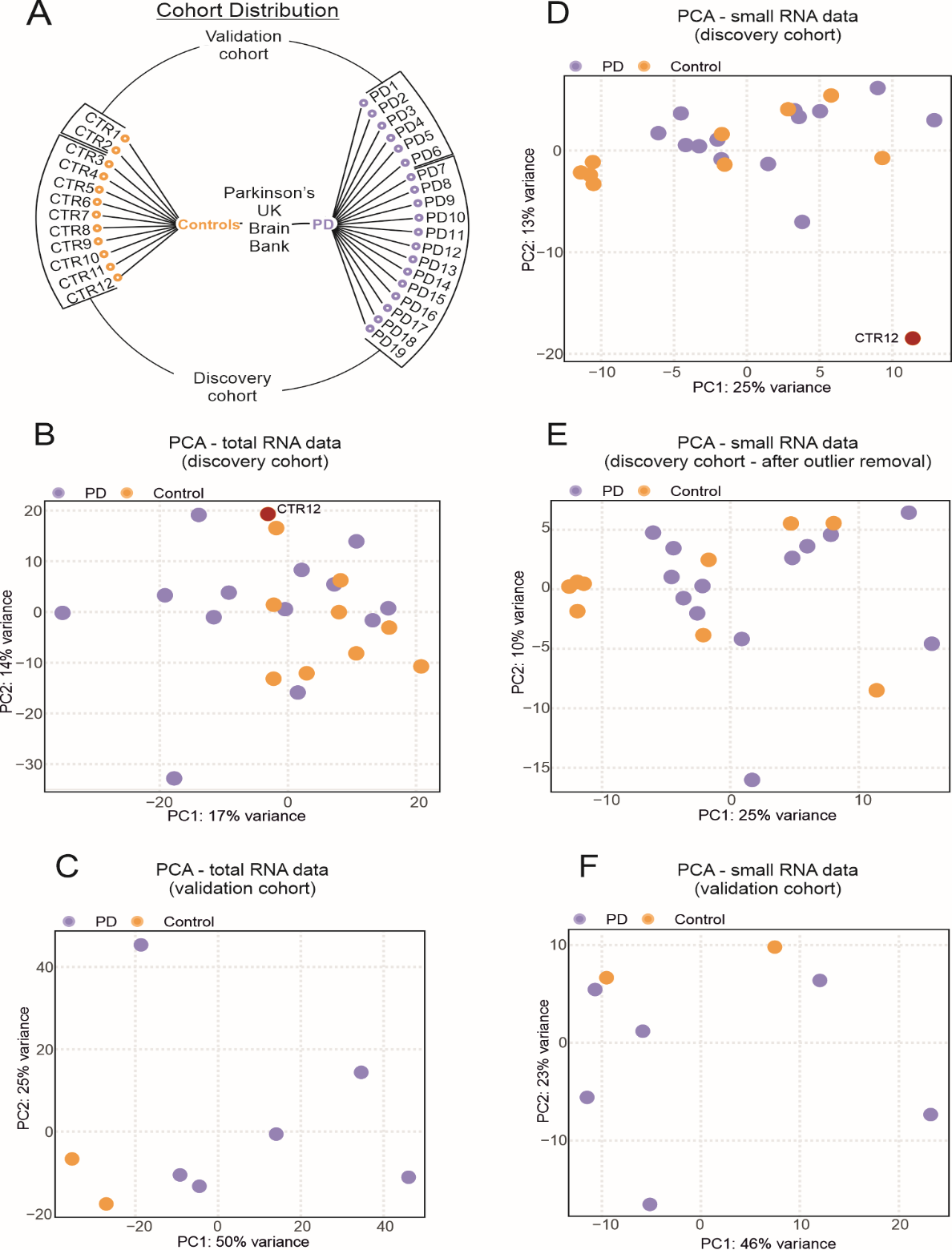

**Figure S3. Cohort composition and Principal Component Analyses (PCA) for RNA Sequencing experiments. (A) Cohort composition for the human postmortem midbrain samples obtained from the Parkinson’s UK Brain Bank. The outlines indicate the samples from the discovery cohort, composed by 13 Parkinson’s disease (PD) patients and 10 controls (CTR), and the validation cohort, which included 6 PD and 2 CTR. (B) PCA for the total RNA data for the discovery cohort, with the control sample CTR12 removed from the analysis for the small RNA-seq data coloured in red. (C) PCA for the total RNA data for the validation cohort. (D) PCA for the small RNA data for the discovery cohort. Outlier sample CTR12 lies separately from the rest of the samples (details in the Results section). (E) PCA for small RNA data for the discovery cohort after outlier removal. (F) PCA for the small RNA data for the validation cohort.**

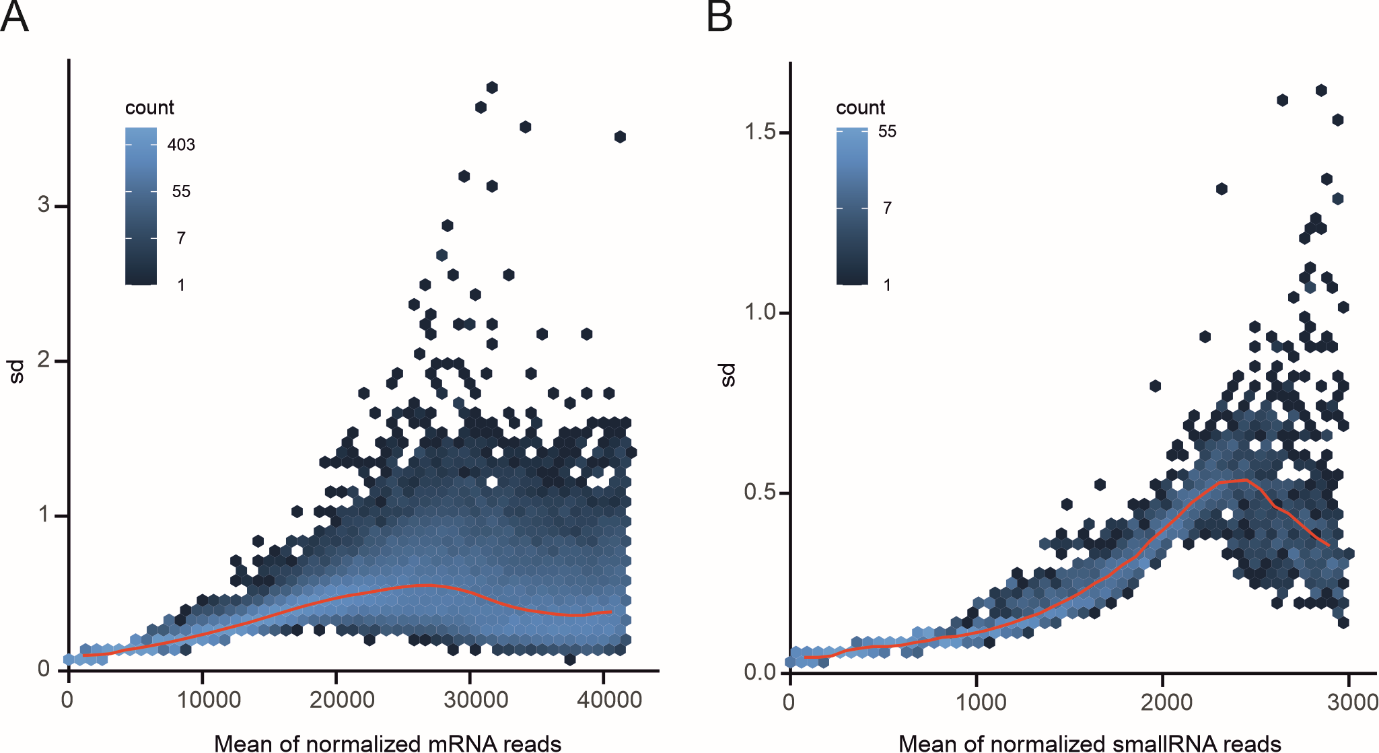

**Figure S4. Mean dispersion estimation plots for (A) mRNA and (B) smallRNA count reads. The y-axis represents the standard deviation (sd) of the mean counts distribution, the color scale indicates the mean count intensity, and the red curve is the fit between dispersion and average through maximum likelihood estimates (MLEs).**

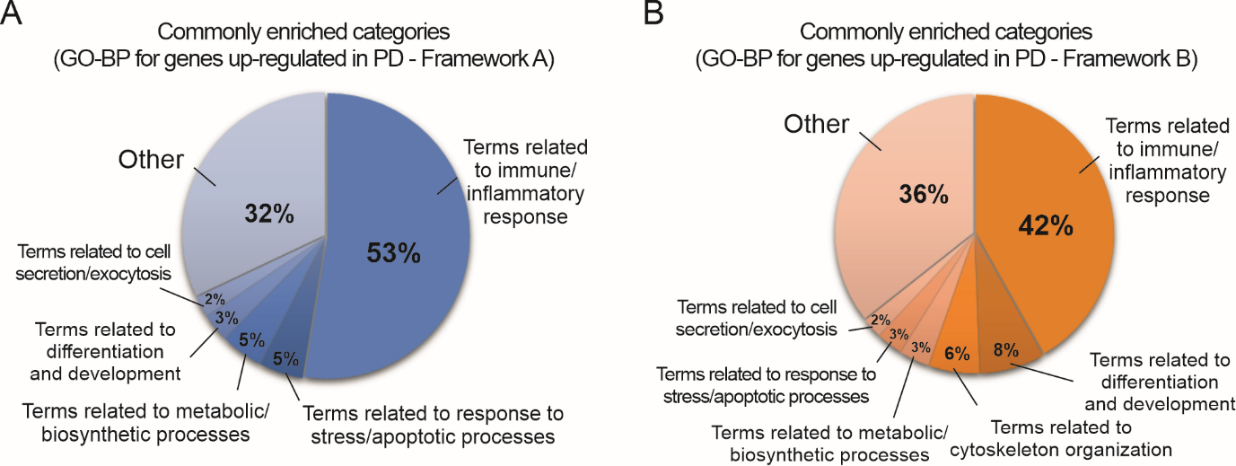

**Figure S5. Functional enrichment analysis (GO-biological processes and KEGG pathways) of differentially up-regulated genes in PD. (A) Summary of enriched GO-*biological processes* (GO-BP) categories in the functional annotation for genes up-regulated in PD obtained with framework “A”. (B) Summary of enriched GO-*biological processes* (GO-BP) categories in the functional annotation for genes up-regulated in PD obtained with framework “B”.**

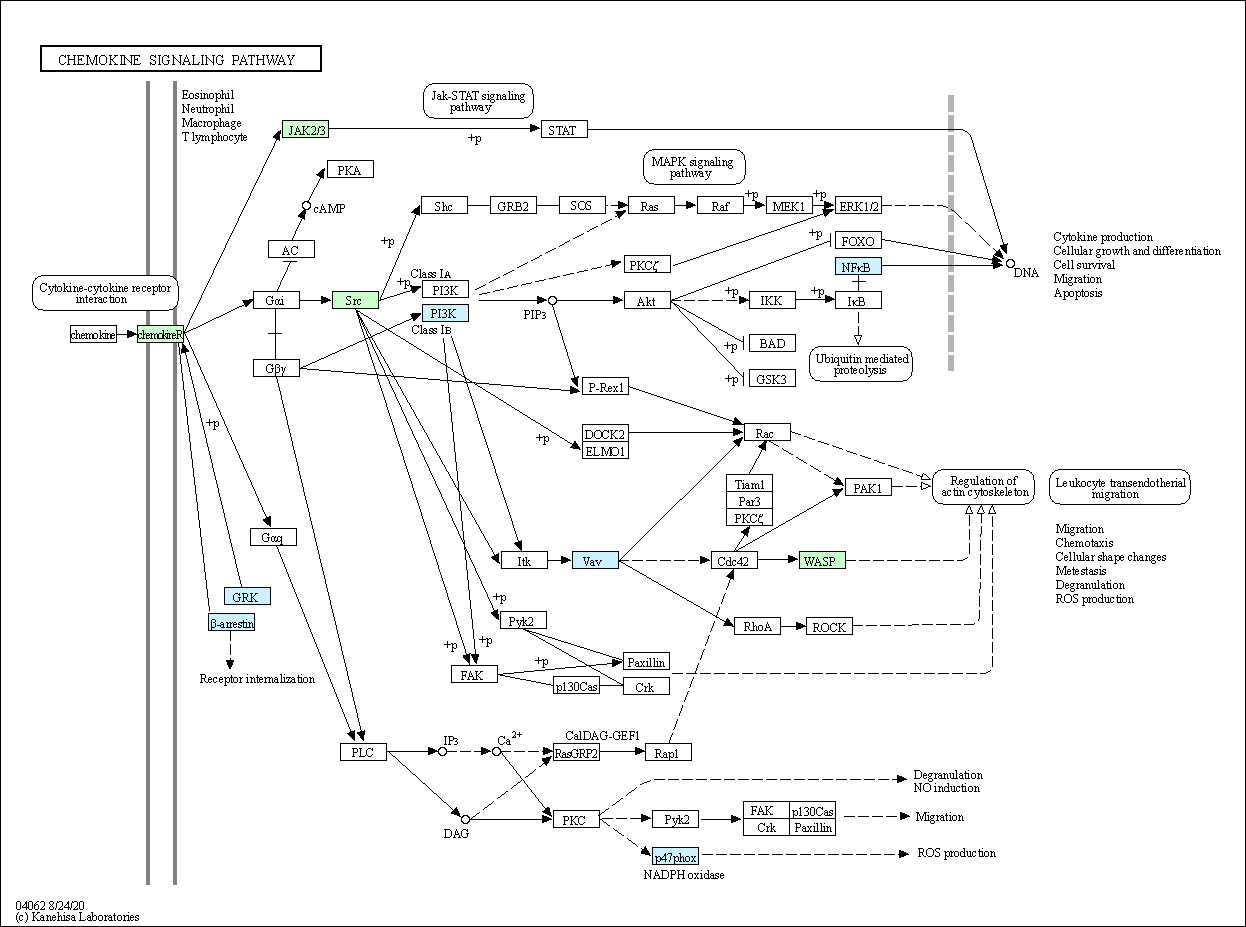
**Figure S6. KEGG human chemokine signaling pathway colored based on the enrichment analysis (FDR < 0.05) of up-regulated significant genes. Green represents the elements statistically enriched in this pathway for both frameworks “A” and “B” and blue only for framework “A”.**

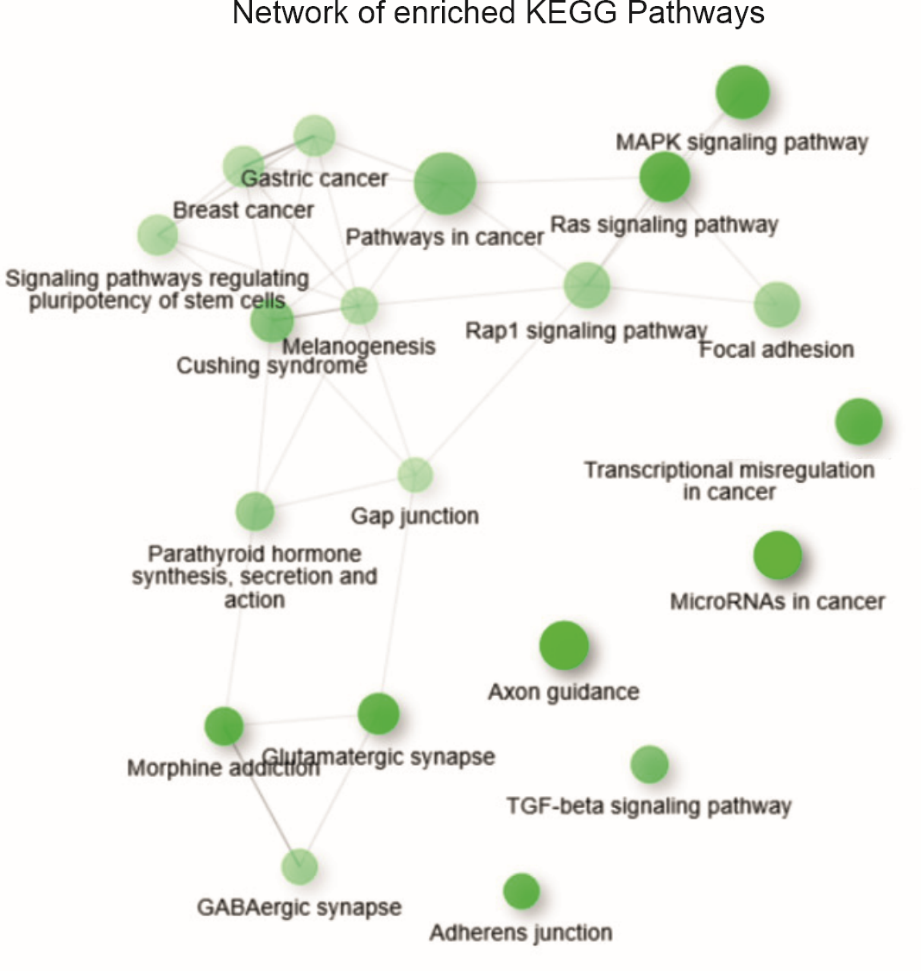

**Figure S7. Network analysis of *KEGG Pathways* for the genes up-regulated in PD with both analysis frameworks. Network shows the relationship between enriched terms provided by the ShinyGO v0.61 platform. Connections between pathways denote 20% or more genes shared. Darkness of nodes coded with the level of significance of the enrichment, while node size is representative for the gene set size. Thicker edges represent more overlapped genes.**

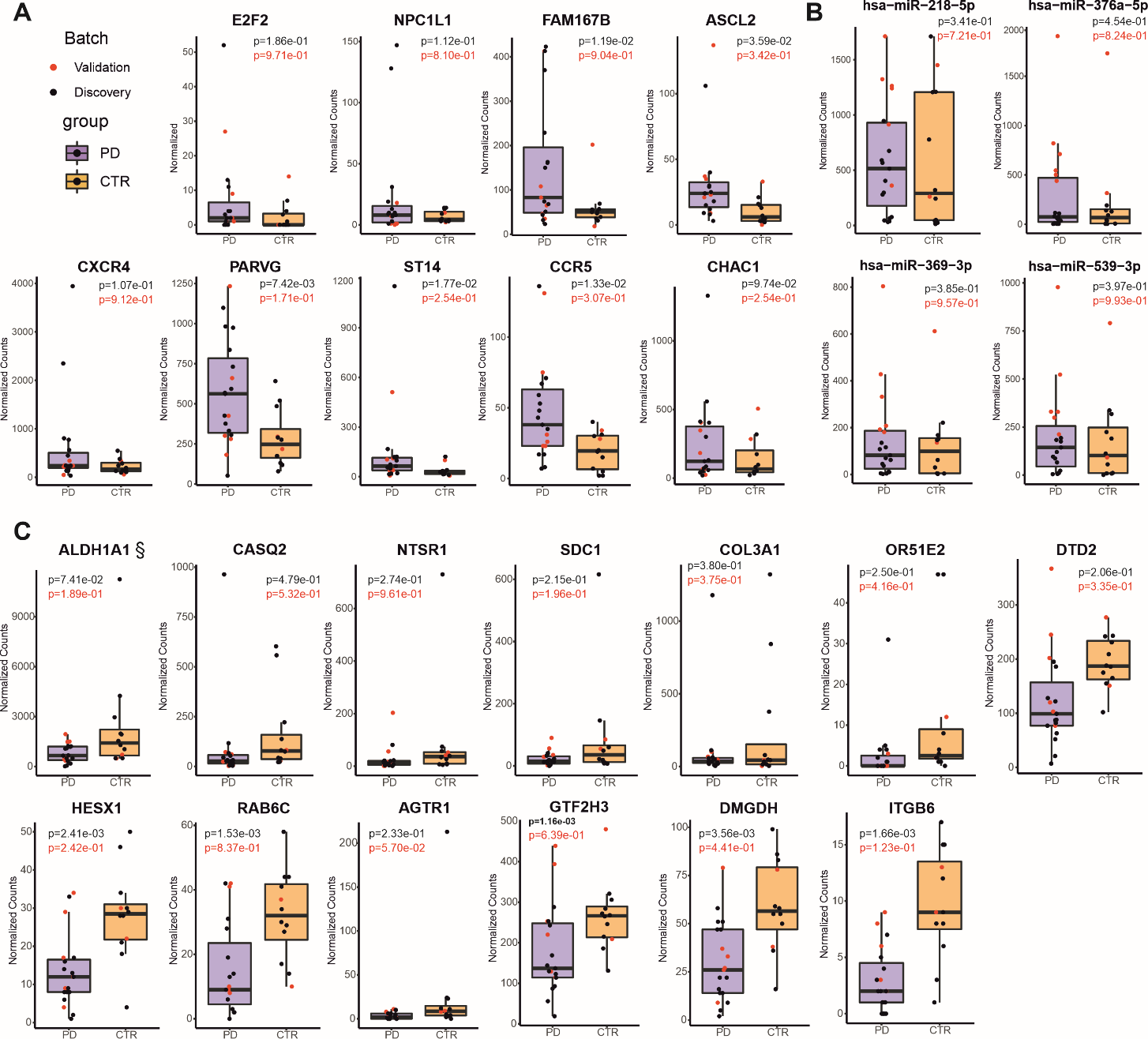

**Figure S8. Barplots of normalized counts for RNA Sequencing experiments, with discovery and validation data cohorts. (A) miRNAs (significant). (B) Up-regulated genes with a respective miRNA pair (found with both frameworks). (C) Down-regulated regulated genes with a respective miRNA pair (found with both frameworks). (§) retinal dehydrogenase 1 (ALDH1A1), to which a valid link was identified after integrating both the miRNA-transcript expression as well as the transcript-protein expression. The p-values were calculated using a two-sided Student’s t-test between PD and CTR samples for each omic analysis.**

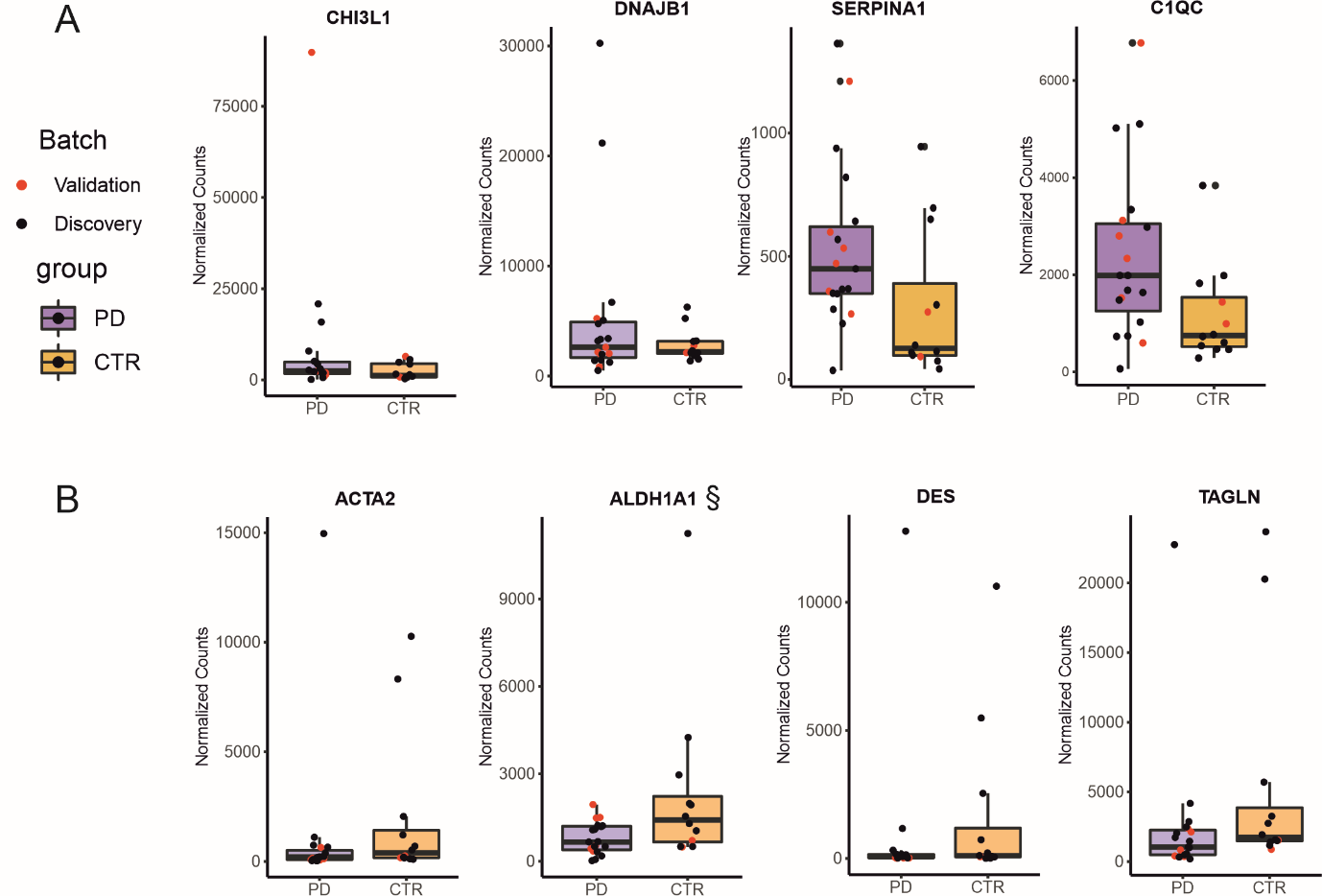

**Figure S9. Transcripts with a valid link in the proteome. (A) Genes up-regulated in PD with a respective protein pair (found in both frameworks). (B) Genes down-regulated in PD with a respective protein pair (found with both frameworks). (§) highlights retinal dehydrogenase 1 (ALDH1A1), to which a valid link was identified after integrating both the miRNA-transcript expression as well as the transcript-protein expression. Data is given in normalized counts, including validation and discovery cohort. The p-values were calculated using a two-sided Student’s t-test between PD and CTR samples for each omic.**

**Supplementary tables**

**Table S1. Genes/Exons assessed in MLPA experiments**

| **Gene/Exon** | **Chromosomal band** | **hg18 location** |
| --- | --- | --- |
| TNFRSF9-2 | 01p36.23 | 01-007,923428 |
| PARK7-1 | 01p36.23 | 01-007,944030 |
| PARK7-1 | 01p36.23 | 01-007,944328 |
| PARK7-2 | 01p36.23 | 01-007,945409 |
| PARK7-3 | 01p36.23 | 01-007,947986 |
| PARK7-4 | 01p36.23 | 01-007,951947 |
| PARK7-5 | 01p36.23 | 01-007,953554 |
| PARK7-6 | 01p36.23 | 01-007,960310 |
| PARK7-7 | 01p36.23 | 01-007,967679 |
| ATP13A2-9 | 01p36.13 | 01-017,199465 |
| ATP13A2-2 | 01p36.13 | 01-017,204821 |
| PINK1-1 | 01p36.12 | 01-020,832858 |
| PINK1-2 | 01p36.12 | 01-020,836975 |
| PINK1-3 | 01p36.12 | 01-020,839042 |
| PINK1-4 | 01p36.12 | 01-020,843555 |
| PINK1-5 | 01p36.12 | 01-020,844754 |
| PINK1-6 | 01p36.12 | 01-020,847671 |
| PINK1-7 | 01p36.12 | 01-020,848135 |
| PINK1-8 | 01p36.12 | 01-020,850116 |
| SNCA-6 | 04q22.1 | 04-090,866748 |
| SNCA-5 | 04q22.1 | 04-090,869382 |
| SNCA-4 | 04q22.1 | 04-090,962507 |
| SNCA-3 | 04q22.1 | 04-090,968331 |
| SNCA-2 (A30P) | 04q22.1 | 04-090,975721 |
| SNCA-2 | 04q22.1 | 04-090,975795 |
| SNCA-1 | 04q22.1 | 04-090,977117 |
| LPA-39 | 06q25.3 | 06-160,873567 |
| PARK2-12 | 06q26 | 06-161,690736 |
| PARK2-11 | 06q26 | 06-161,701126 |
| PARK2-10 | 06q26 | 06-161,727843 |
| PARK2-9 | 06q26 | 06-161,889882 |
| PARK2-8 | 06q26 | 06-161,910375 |
| PARK2-7 | 06q26 | 06-162,126840 |
| PARK2-6 | 06q26 | 06-162,314327 |
| PARK2-5 | 06q26 | 06-162,395114 |
| PARK2-4 | 06q26 | 06-162,542097 |
| PARK2-3 | 06q26 | 06-162,603527 |
| PARK2-2 | 06q26 | 06-162,784395 |
| PARK2-1 | 06q26 | 06-163,068646 |
| LRRK2-41 (G2019S) | 12q12 | 12-039,020438 |
| Reference | 02p13.2 | 02-071,750350 |
| Reference | 02q36.3 | 02-227,881969 |
| Reference | 03p22.2 | 03-038,494904 |
| Reference | 05q31.1 | 05-132,037610 |
| Reference | 06p12.3 | 06-051,605391 |
| Reference | 08q12.2 | 08-061,883322 |
| Reference | 09p13.3 | 09-034,483216 |
| Reference | 11p14.3 | 11-022,251032 |
| Reference | 15q21.1 | 15-042,674827 |
| Reference | 18q21.1 | 18-045,743292 |

**Table S2. List of genes assessed in Gene panel sequencing experiments**

Targeted Sequencing (Gene Panel) - Evaluated Genes

*ADCY5*

*ANO3*

*COX20*

*DJ-1*

*GBA*

*GCH1*

*GNAL*

*KMT2B*

*LRRK2*

*MCOLN1*

*Parkin*

*PDGFB*

*PDGFRB*

*PINK1*

*PLA2G6*

*POLG*

*PRKRA*

*RAB12*

*RAB39B*

*SGCE*

*SLC20A2*

*SNCA*

*TAF1 (4 variants)*

*THAP1*

*TOR1A*

*VAC14*

*VPS13C*

*VPS35*

*XPR1*

**Table S3. RNA-Seq decomposition score results using xCell and MCP-counter methods.** Log_2_(Fold-Change) (Log_2_FC) = score_PD_ - score_CTR_, p-values obtained using a Wilcoxon t-test and adjusted by Bonferroni's correction method.

| **xCell** | | | |
| --- | --- | --- | --- |
| **Cell Type** | **Log_2_FC** | **p-value** | **p-adjusted value** |
| **Myeloid dendritic cell activated** | -2.185 | 9.483e-02 | 3.082e-01 |
| **T cell CD4+ (non-regulatory)** | 1.499 | 7.950e-02 | 2.819e-01 |
| **Granulocyte-monocyte progenitor** | 51.581 | 6.194e-02 | 2.416e-01 |
| **Class-switched memory B cell** | -1.895 | 5.863e-02 | 2.416e-01 |
| **T cell CD4+ central memory** | -2.355 | 5.393e-02 | 2.416e-01 |
| **B cell naive** | 3.711 | 4.430e-02 | 2.416e-01 |
| **T cell regulatory (Tregs)** | 3.359 | 2.968e-02 | 1.929e-01 |
| **Endothelial cell** | 2.607 | 2.696e-02 | 1.929e-01 |
| **B cell memory** | 2.512 | 2.324e-02 | 1.929e-01 |
| **Common myeloid progenitor** | -49.819 | 2.308e-02 | 1.929e-01 |
| **T cell CD4+ memory** | 4.524 | 1.429e-02 | 1.929e-01 |
| **Hematopoietic stem cell** | 3.566 | 7.761e-06 | 3.027e-04 |
| **MCP-counter** | | | |
| **Cell Type** | **Log_2_FC** | **p-value** | **p-adjusted value** |
| **T cell** | -6.025e-01 | 9.424e-02 | 5.183e-01 |
| **cytotoxicity score** | 1.021 | 8.350e-02 | 5.183e-01 |

**Table S4. Integration pairs between significantly DE miRNAs and predicted/validated mRNA/gene targets from framework B.**

| **Validated targets - mirTarBase** | | | | | | | |
| --- | --- | --- | --- | --- | --- | --- | --- |
| **Gene Name** | **miRNA** | **log_2_FC miRNA** | **p-value miRNA** | **padj miRNA** | **log_2_FC gene** | **p-value gene** | **padj gene** |
| CREBBP | hsa-miR-218-5p | 6.366e-01 | 2.394e-04 | 9.122e-02 | 3.513e-01 | 3.161e-04 | 9.181e-02 |
| PAPD7 | hsa-miR-218-5p | 6.366e-01 | 2.394e-04 | 9.122e-02 | 3.802e-01 | 5.215e-04 | 9.934e-02 |
| POU2F2 | hsa-miR-218-5p | 6.366e-01 | 2.394e-04 | 9.122e-02 | 8.656e-01 | 3.785e-04 | 9.181e-02 |
| RBM6 | hsa-miR-218-5p | 6.366e-01 | 2.394e-04 | 9.122e-02 | 4.343e-01 | 4.046e-04 | 9.181e-02 |
| **Predicted targets - miRDB** | | | | | | | |
| **Gene Name** | **miRNA** | **log_2_FC miRNA** | **p-value miRNA** | **padj miRNA** | **log_2_FC gene** | **p-value gene** | **padj gene** |
| ETV5 | hsa-miR-369-3p | 6.984e-01 | 1.862e-04 | 9.122e-02 | 7.051e-01 | 5.476e-04 | 9.934e-02 |
| GTF2H3 | hsa-miR-369-3p | 6.984e-01 | 1.862e-04 | 9.122e-02 | -4.633e-01 | 2.225e-04 | 4.081e-02 |
| JADE2 | hsa-miR-218-5p | 6.366e-01 | 2.394e-04 | 9.122e-02 | 6.022e-01 | 4.012e-04 | 9.181e-02 |
| RAB6C | hsa-miR-218-5p | 6.366e-01 | 2.394e-04 | 9.122e-02 | -8.860e-01 | 3.331e-04 | 9.181e-02 |
| RCOR1 | hsa-miR-218-5p | 6.366e-01 | 2.394e-04 | 9.122e-02 | 3.516e-01 | 6.048e-04 | 9.933e-02 |
| SLC38A2 | hsa-miR-369-3p | 6.984e-01 | 1.862e-04 | 9.122e-02 | 9.138e-01 | 1.561e-04 | 7.404e-02 |
